# Neutrophil-fibroblast crosstalk drives immunofibrosis in Crohn’s disease through IFNα pathway

**DOI:** 10.1101/2023.09.08.23295281

**Authors:** Efstratios Gavriilidis, Georgios Divolis, Anastasia-Maria Natsi, Nikolaos Kafalis, Dionysios Kogias, Christina Antoniadou, Evgenia Synolaki, Evgenios Pavlos, Marianna A. Koutsi, Stylianos Didaskalou, Victoria Tsironidou, Ariana Gavriil, Vasileios Papadopoulos, Marios Agelopoulos, Dimitrios Tsilingiris, Maria Koffa, Alexandra Giatromanolaki, Georgios Kouklakis, Konstantinos Ritis, Panagiotis Skendros

**Author notes:** These authors contributed equally to this work and share first authorship (Efstratios Gavriilidis, Georgios Divolis, Anastasia-Maria Natsi). Address correspondence to: Konstantinos Ritis, Laboratory of Molecular Hematology, Department of Medicine, Democritus University of Thrace, Alexandroupolis, 68100, Greece.; Panagiotis Skendros, First Department of Internal Medicine, University Hospital of Alexandroupolis, Democritus University of Thrace, Alexandroupolis, 68100, Greece.

## Abstract

This study investigated the interaction between neutrophils and intestinal fibroblasts in Crohn’s disease (CD) immunofibrosis. Peripheral neutrophils, enriched-neutrophil extracellular traps (eNETs), serum, primary intestinal fibroblasts (PIFs) and intestinal biopsies were studied. Neutrophils’ RNA-sequencing, multi-cytokine profiling and cell-based functional assays at mRNA/protein level were performed. Compared to ulcerative colitis (UC), PIFs from CD patients displayed a distinct fibrotic phenotype characterized by negative Krüppel-like Factor-2 (KLF2) and increased cellular communication network factor-2 (CCN2) expression leading to collagen production. PIFs-derived IL-8 appears as a culprit chemoattractant of neutrophils in the intestine, where CD neutrophils were accumulated close to fibrotic lesions. Functionally, only CD neutrophils *via* eNETs can induce a CD-like phenotype in HI PIFs. High serum IFNα and IFΝ-responsive signature in neutrophils were observed in CD, distinguishing it from UC. Moreover, CD serum can stimulate the release of fibrogenic eNETs in an IFNα-dependent manner, suggesting the priming role of IFNα in circulating neutrophils. Inhibition of eNETs or JAK signaling in neutrophils or PIFs prevented the neutrophil-mediated fibrotic effect on PIFs. Furthermore, serum IFNα and transcripts of key IFN-signaling components in neutrophils were well-correlated with CD severity. This study reveals the role of IFNα/neutrophil/fibroblast axis in CD immunofibrosis, suggesting candidate biomarkers and therapeutic targets.

## 1. INTRODUCTION

Crohn’s disease (CD) is characterized by chronic relapsing inflammation that may lead to intestinal fibrosis causing lifelong disabling illness with a large impact on the quality of life and the healthcare systems (1, 2). Therefore, understanding the mechanisms of initiation and propagation of intestinal fibrosis in CD is crucial to providing knowledge for diagnosis and better care of patients (3, 4).

Cellular plasticity is fundamental to human immunity and, as recently recognized, a key aspect of neutrophil biology (5, 6). Recent studies suggest that circulating neutrophils, as an adaptation to the different environmental conditions, undergo transcriptional reprogramming that allows them to acquire disease-specific phenotypes and commit their cell-fate plasticity upon their entrance to the site of tissue inflammation attracted by tissue-derived chemotactic factors (6). Activated neutrophils release a plethora of antimicrobial and proinflammatory mediators on extracellular vesicles and traps that could dictate their diverse functional role in different diseases (7–9).

In line with these, previous studies indicated that activated neutrophils through the release of neutrophil extracellular traps (NETs) may play a pro-inflammatory or fibrotic role by promoting the differentiation and activation of human fibroblasts (10–12). More recently, our group suggested that NETs and downregulation of transcriptional factor Krüppel-like Factor 2 (KLF2) in human lung fibroblasts are linked with the inflammatory environment of COVID-19 that leads to immunofibrosis (13). Whether neutrophils exert immunofibrotic effects on intestinal fibroblasts and how these cells interact with each other in the inflammatory environment of CD leading to tissue damage is largely unknown yet (4, 14).

This study provides a new understanding of the mechanisms involved in CD immunofibrosis through neutrophils/NETs functional plasticity. We identified that, in contrast to UC, neutrophils of CD are primed by IFNα/JAK signaling to commit an active role on intestinal fibroblasts, thus inducing their distinct fibrotic phenotype. The production of IL-8 by intestinal fibroblasts sustains their mutual interaction with neutrophils. Translating these findings, the levels of key IFN type I pathway components are positively correlated with disease severity in CD patients promising novel diagnostic and therapeutic targets.

## 2. RESULTS

### 2.1 Fibroblasts of Crohn’s disease patients display a distinct fibrotic phenotype

Since fibrotic complications are a prominent feature of Crohn’s disease (CD) (2, 3) and recent data have shown that profibrotic activity of human lung fibroblasts is associated with downregulated KLF2, elevated CCN2 levels, and increased collagen production (13), we aimed to investigate whether intestinal fibroblasts in CD share similarities with this fibrotic phenotype.

We studied the phenotype of cultured primary intestinal fibroblasts (PIFs), isolated from treatment-naïve patients with active CD and UC, as well as from HI. UC has been included in this study as a control inflammatory bowel, non-typical fibrotic, disease. In contrast to UC, CD PIFs, compared to HI PIFs, showed significantly lower KLF2 and higher CCN2 mRNA and protein levels (**Fig. 1A-D**), as well as increased collagen release (**Fig. 1E**).

**Figure 1.**
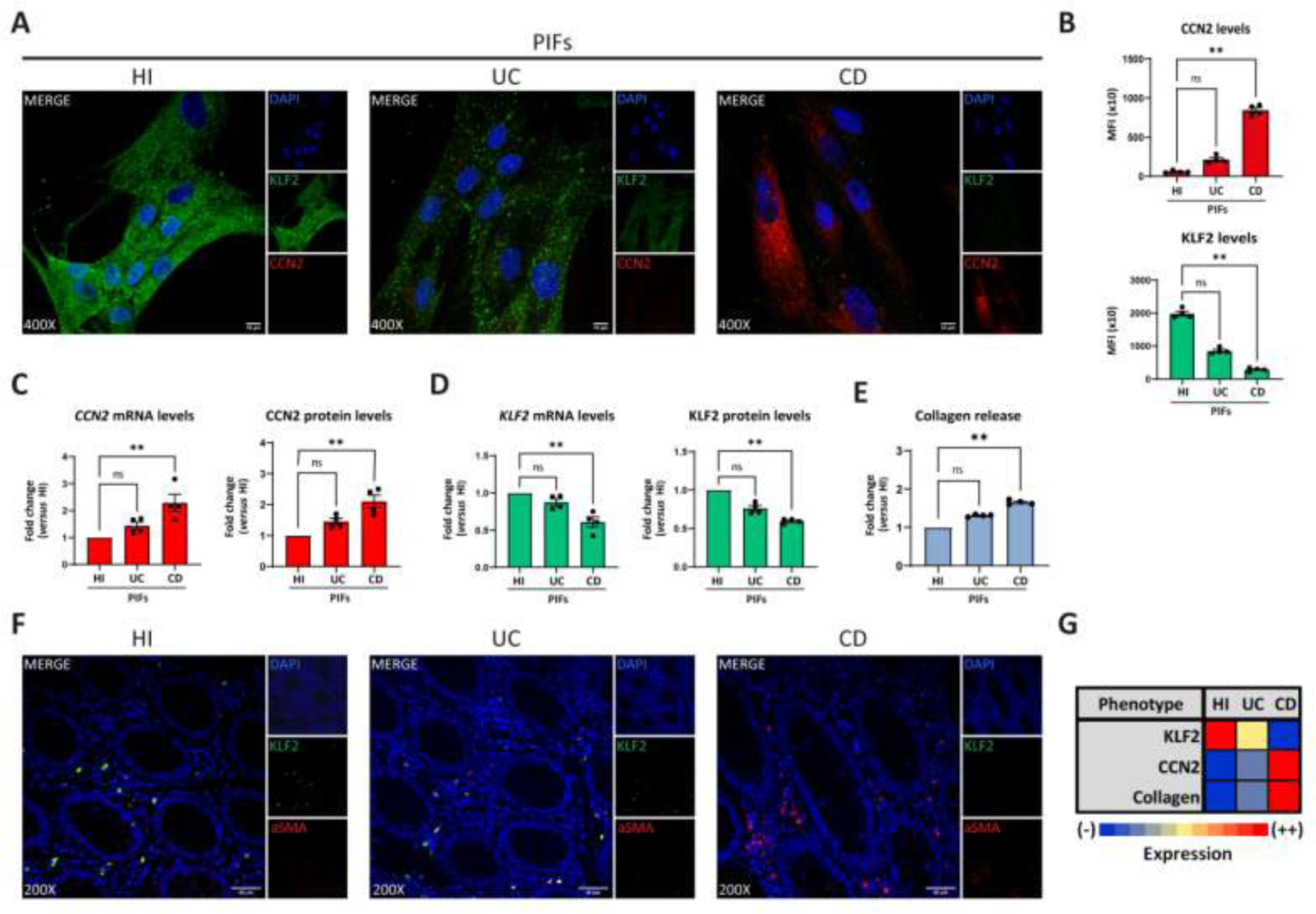
Crohn’s disease intestinal fibroblasts are characterized by a distinct fibrotic phenotype. Assessment of KLF2 and CCN2 expression in PIFs from IBD patients and HI by **(A)** immunostaining (blue: DAPI, green: KLF2, red: CCN2), and **(B)** corresponding MFI quantification. **(C)** CCN2 and **(D)** KLF2 mRNA and protein levels were also assessed by qPCR and in-cell ELISA. **(E)** Collagen release was measured in supernatants collected from the above-mentioned PIFs. **(F)** KLF2 expression in myofibroblasts within intestinal biopsies obtained from the same patients (blue: DAPI, green: KLF2, red: aSMA). **(G)** Heatmap indicating the expression intensity of KLF2, CCN2 and collagen release in PIFs. **(A,F)** One representative example out of four independent experiments is shown. Confocal microscopy. **(Α)** Magnification: 400x, Scale Bar: 10μm **(F)** Magnification: 200x, Scale Bar: 40μm. Nonparametric Kruskal-Wallis followed by Dunn’s multiple comparisons test was applied in all panels, *n=4*, **p<0.01, n.s.: not significant. Data are expressed as mean ± SEM. aSMA, anti-smooth muscle antibody; CD, Crohn’s disease; CCN2, cellular communication network factor 2; HI, healthy individuals; IBD, inflammatory bowel disease; KLF2, Kruppel-like factor 2; MFI, mean fluorescence intensity; PIFs, primary intestinal fibroblasts; UC, ulcerative colitis.

Next, we examined intestinal tissues obtained from the same patients. An abundance of aSMA positive fibroblasts, was observed in CD compared to UC tissues. These CD myofibroblasts showed absence of KLF2 staining in contrast to KLF2-positive cells in UC (**Fig. 1F**). Similar to isolated PIFs, in tissues stained for vimentin, CD fibroblasts were also KLF2 negative, while UC and HI tissue fibroblasts showed faint and bright KLF2 staining, respectively (**Fig. S1A-C**).

Collectively, we observed a negative association between KLF2 and CCN2 in the fibroblasts of CD patients. CD fibroblasts expressed the highest mRNA and protein levels of CCN2 in association with the highest collagen production, whereas HI fibroblasts expressed the highest levels of KLF2 with concomitant absence of CCN2 expression and collagen production. Considering these, we suggested an arbitrary grading scale, according to the expression intensity of the abovementioned parameters (**Fig. 1G**). CD PIFs demonstrated fibrotic activity and displayed a distinct phenotype, compared to UC PIFs. For the sake of brevity, we characterized the former as KLF2 (-), CCN2 (++), collagen (++) cells, in contrast to UC KLF2 (+), CCN2 (-), collagen (-) PIFs and HI KLF2 (++), CCN2 (-), collagen (-) PIFs (**Fig. 1G**).

### 2.2 The fibroblast-derived IL-8 is associated with the presence of neutrophils in the intestine of patients with inflammatory bowel disease

Since previous studies suggested that neutrophils may activate fibroblasts and interfere in the fibrotic process (10–12), we sought to investigate the distribution of neutrophils in intestinal tissues of active IBD, as well as their association with fibrotic lesions, in the same patients described above. Higher numbers of neutrophils were observed in UC, compared to CD intestinal tissues. (**Fig. 2A**). However, neutrophils in intestinal sections of CD were found in the vicinity of the fibrotic areas, as defined by positive Masson’s trichrome staining for collagen, a finding that was not observed in UC tissues (**Fig. 2A,B**). Neutrophils and fibrosis, as expected, were absent from healthy intestinal tissues (**Fig. 2A,B**)

**Figure 2.**
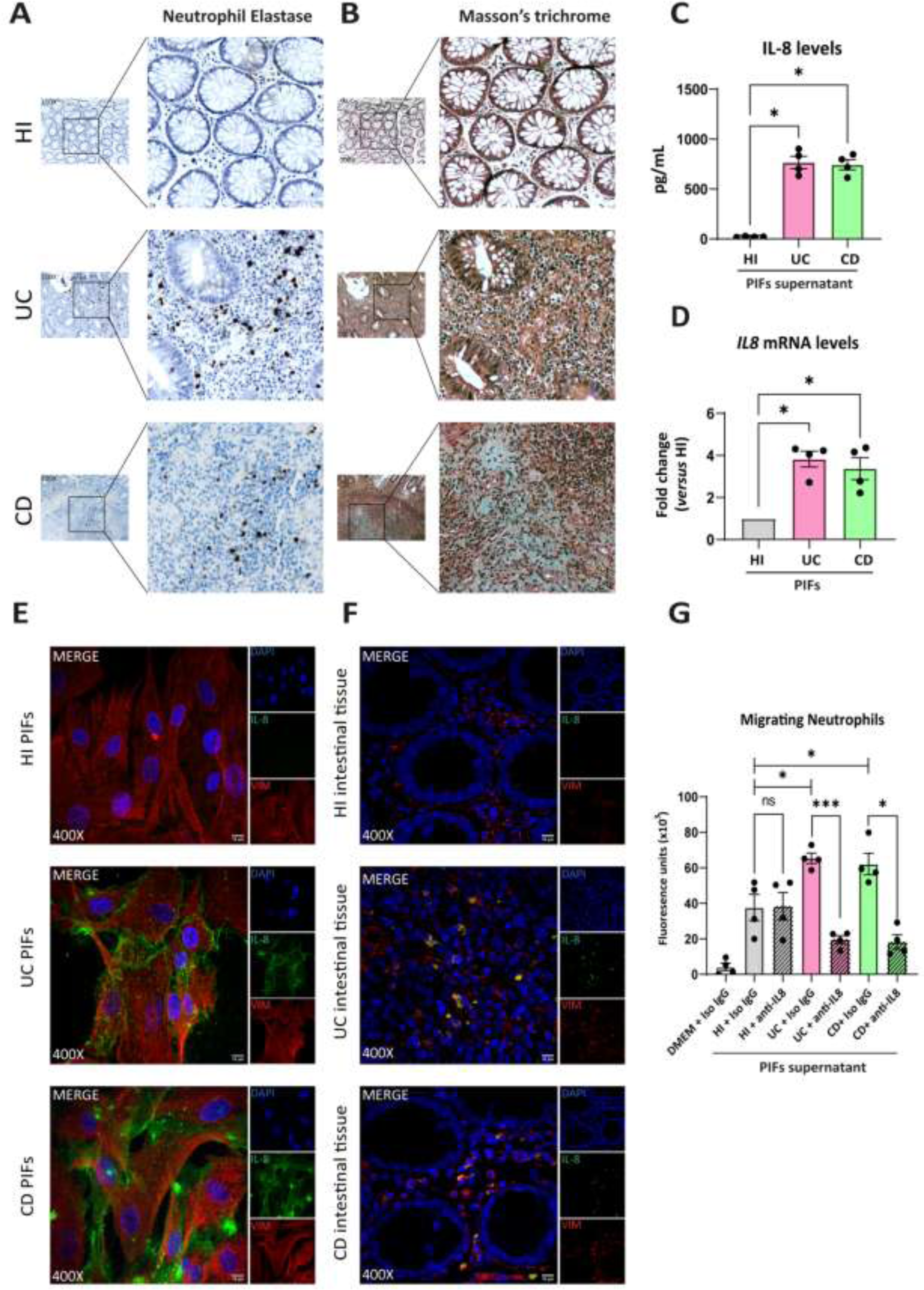
UC and CD intestinal tissues are characterized by differential spatial distribution of neutrophils, attracted in the intestinal environment by fibroblast-derived IL-8. **(A)** Neutrophil Elastase IHC staining (brown cells) and **(B)** Masson’s trichrome (cyan fibers) indicating the presence of neutrophils and fibrotic areas respectively, in serial cross sections obtained from the same intestinal biopsies. Thickness between the serial cross sections in **(A)** and **(B)** was 4 μm. **(C)** IL-8 levels in supernatants of PIFs measured by a bead-based flow cytometric assay. **(D)** IL-8 mRNA as assessed by qPCR and **(E)** protein levels in PIFs immunostaining (blue: DAPI, green: IL-8, red: Vimentin). **(F)** IL-8 expression by immunostaining in intestinal tissue fibroblasts (blue: DAPI, green: IL-8, red: Vimentin). **(G)** Chemotactic capacity of the PIFs’ supernatant on HI neutrophils, before and after the neutralization of IL-8, as assessed by a transwell migration assay. **(A,B,E,F)** One representative example out of four independent experiments is shown. **(A,B)** Optical microscopy, magnification: 100x, **(E,F)** Confocal microscopy, magnification: 400x, Scale Bar: 10μm. Nonparametric Kruskal-Wallis followed by Dunn’s multiple comparisons test was performed in **(C)** and (**D)**, *n=4*, *p<0,05, n.s.: not significant. **(G)** Bayesian unpaired t-tests followed by the Benjamini-Hochberg correction, were used to compare the migratory capacity of HI PIFs supernatants to UC and CD. For comparisons between PIFs supernatants that were treated with IL-8 neutralizing antibody (anti-IL-8), and supernatants treated with IgG isotype control (Iso IgG), Bayesian paired t-tests were performed, *n=4*, *p<0,05, ***p<0.001, n.s.: not significant. Data are expressed as mean ± SEM. CD, Crohn’s disease; DMEM, Dulbecco’s Modified Eagle Medium; HI, healthy individuals; IHC, immunohistochemistry; PIFs, primary intestinal fibroblasts; UC, ulcerative colitis;

Next, to decipher the mechanism by which neutrophils become attracted to the intestinal environment, we investigated whether fibroblasts produce known chemoattractant factors. Thus, we found that IL-8 (CXCL8) was the chemokine with significantly higher levels in supernatants of cultured PIFs obtained from UC and CD compared to HI (**Fig. 2C**). Increased IL-8 expression was also observed in isolated PIFs both at mRNA and protein levels, as well as in fibroblasts of intestinal sections (**Fig. 2D-F**). No significant alterations were observed in the levels of other cytokines (**Fig. S2**).

Prompted by these results and in view that IL-8 has a distinct target specificity for neutrophils (15), we next performed a chemotactic assay that indicated increased migratory capacity of neutrophils when stimulated by the IL-8-rich supernatant of UC and CD PIFs, an effect that was inhibited after IL-8 neutralization (**Fig. 2G**).

These data suggest that both UC and CD fibroblasts express functional IL-8 as a key neutrophil chemoattractant factor of the intestinal environment. However, it warrants further investigation if neutrophils in each disease have differential functional properties during their crosstalk with fibroblasts.

### 2.3 Healthy PIFs acquire a CD-like fibrotic phenotype after treatment with *ex-vivo* enriched neutrophil extracellular traps (eNETs) from CD patients

To investigate the hypothesis that neutrophils of CD, UC patients, or HI have distinct functional properties in their crosstalk with fibroblasts, we obtained mixtures consisting of *ex-vivo* NETs enriched with the supernatant formed during the isolation procedure of NETs, to preserve the total inflammatory environment including structures of DNA scaffold and cellular extracts (eNETs). Moreover, eNETs were collected after *in-vitro* stimulation of HI neutrophils with PMA, a chemical inducer of NETosis, and used as a non-disease-specific stimulus.

Next, we used these neutrophilic mixtures for stimulations on HI PIFs to assess KLF2 and CCN2 mRNA and protein levels, as well as collagen release. We found that HI PIFs stimulated with CD eNETs acquired a CD-like KLF2 (-), CCN2 (++), and collagen (++) fibrotic phenotype (**Fig. 3A,B, G-K**). This effect was not observed in stimulations with HI eNETs (**Fig. 3C, G-K**). In contrast, UC eNETs drive fibroblasts towards the UC-like phenotype, indicating KLF2 (+), CCN2 (-), and collagen (-) cells (**Fig. 3D, G-K**). Moreover, PMA generated mixtures that led fibroblasts towards KLF2 (+), CCN2 (+), and collagen (+) phenotype, indicative of mild fibrotic activity (**Fig. 3E, G-K**). The fact that the serum of CD patients was unable on its own to transform the phenotype of HI fibroblasts towards acquiring fibrotic function, further supports the key role of neutrophils in the activation of fibroblasts (**Fig. 3F-J**).

**Figure 3.**
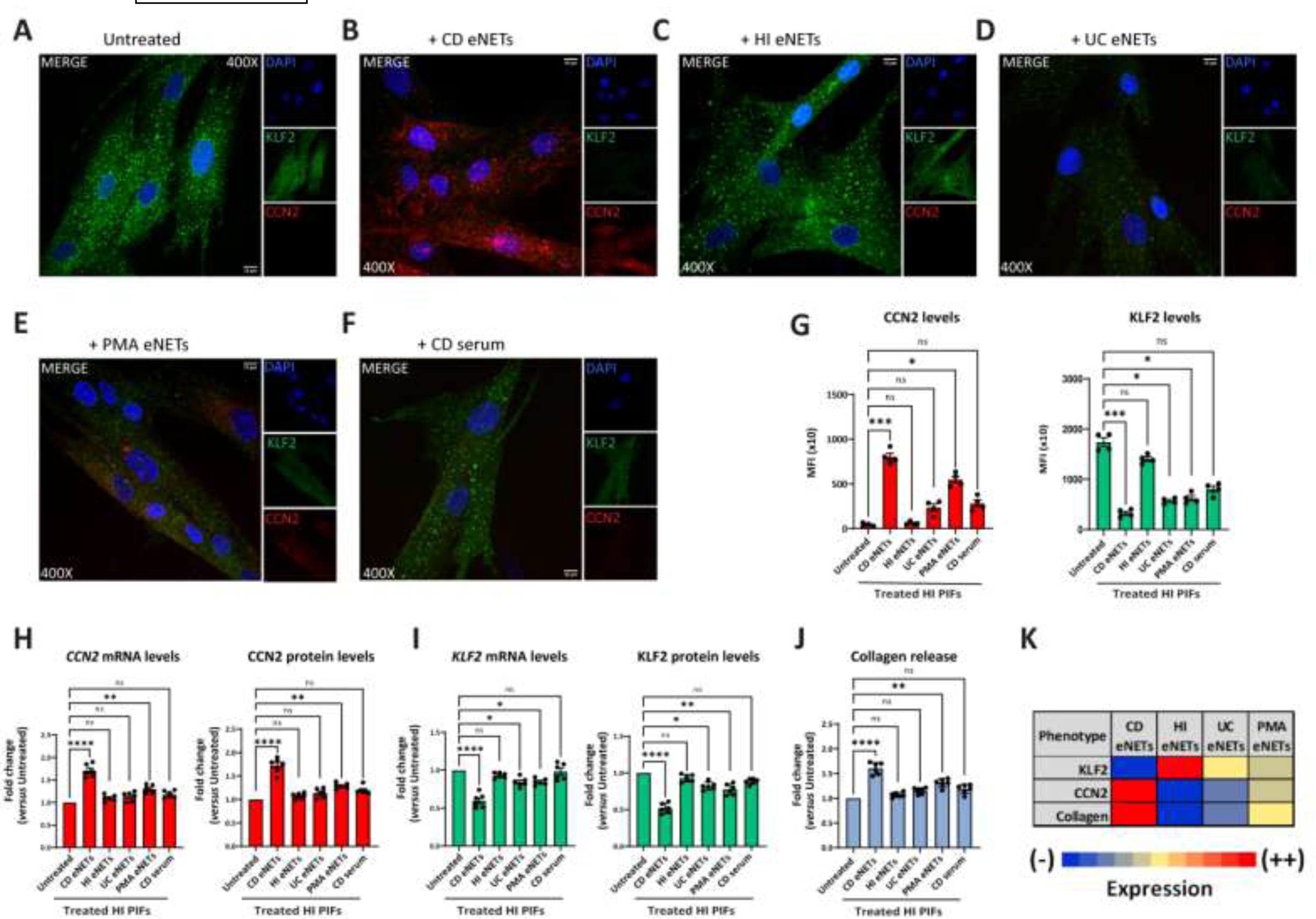
Treatment of healthy PIFs with *ex-vivo* isolated CD eNETs leads to a CD-like fibrotic phenotype. **(A-F)** Immunostaining (blue: DAPI, green: KLF2, red: CCN2) and **(G)** corresponding MFI quantification of HI PIFs treated with *ex-vivo* isolated eNETs from **(B)** CD, **(C)** HI, or **(D)** UC patients, **(E)** PMA-generated eNETs and **(F)** CD serum. The abovementioned treated-PIFs were assessed for CCN2 **(H)** and KLF2 **(I)** mRNA and protein levels by qPCR and in-cell ELISA, respectively, as well as collagen release **(J)**. **(K)** Heatmap depicting the expression intensity of KLF2, CCN2 and collagen release. **(A-F)** One representative example out of four independent experiments is shown. Confocal microscopy. Magnification: 400x, Scale Bar: 10μm. **(G-J)** Nonparametric Kruskal-Wallis followed by Dunn’s multiple comparisons test, *n=6,* *p<0,05, **p<0,01, ***p<0,001, ****p<0,0001, n.s.: not significant. Data are expressed as mean ± SEM. CCN2, cellular communication network factor 2; CD, Crohn’s disease; eNETs, enriched neutrophil extracellular traps; HI, healthy individuals; KLF2, Kruppel-like factor 2; MFI, mean fluorescence intensity; PIFs, primary intestinal fibroblasts; PMA, Phorbol-12-myristate-13-acetate; UC, ulcerative colitis.

Taken together, CD neutrophils probably acquire different plasticity compared to UC neutrophils, being able to transform healthy PIFs to a CD-like fibrotic phenotype, demonstrating a functional role in their crosstalk with intestinal fibroblasts.

### 2.4 Transcriptome analysis of peripheral blood neutrophils unravels distinct pathways in Crohn’s disease and ulcerative colitis

To provide some mechanistic explanation for the differential plasticity observed in IBD neutrophils, we sought to compare the transcriptome of peripheral blood neutrophils isolated from CD and UC patients. Our analysis identified 849 significantly upregulated and 789 downregulated genes in CD neutrophils, whereas 1421 genes were found upregulated and 1110 downregulated in UC neutrophils, relative to control neutrophils purified from healthy individuals (**Fig. 4A**). Interestingly, the two datasets exhibited a substantial overlap since 66% of upregulated and 57% of downregulated genes in CD were also significantly regulated in UC neutrophils (**Fig. 4A, Fig. S3**). Bioinformatics analysis using the GeneCodis4 web-based tool revealed that common upregulated DEGs clustered mainly in immune-related pathways, including neutrophil degranulation, class I MHC mediated processing and presentation, and Toll-like receptor cascades, while translation, mRNA and rRNA processing were amongst the top downregulated processes (**Fig. 4B**). Apart from the commonly regulated DEGs and pathways, we found several genes and processes uniquely regulated in each disease (**Fig. 4B, Fig. S3**). More specifically, interferon signaling was the top pathway selectively upregulated in Crohn’s (**Fig. 4B**), while the majority of unique upregulated genes in UC were involved in the neutrophil degranulation pathway. Regarding the unique downregulated genes, these are clustered mainly to translation- and post-translational modification-related pathways in CD neutrophils, while the respective DEGs in UC neutrophils belonged to the pathways of chromatin organization and apoptotic process among others (**Fig. 4B**).

**Figure 4.**
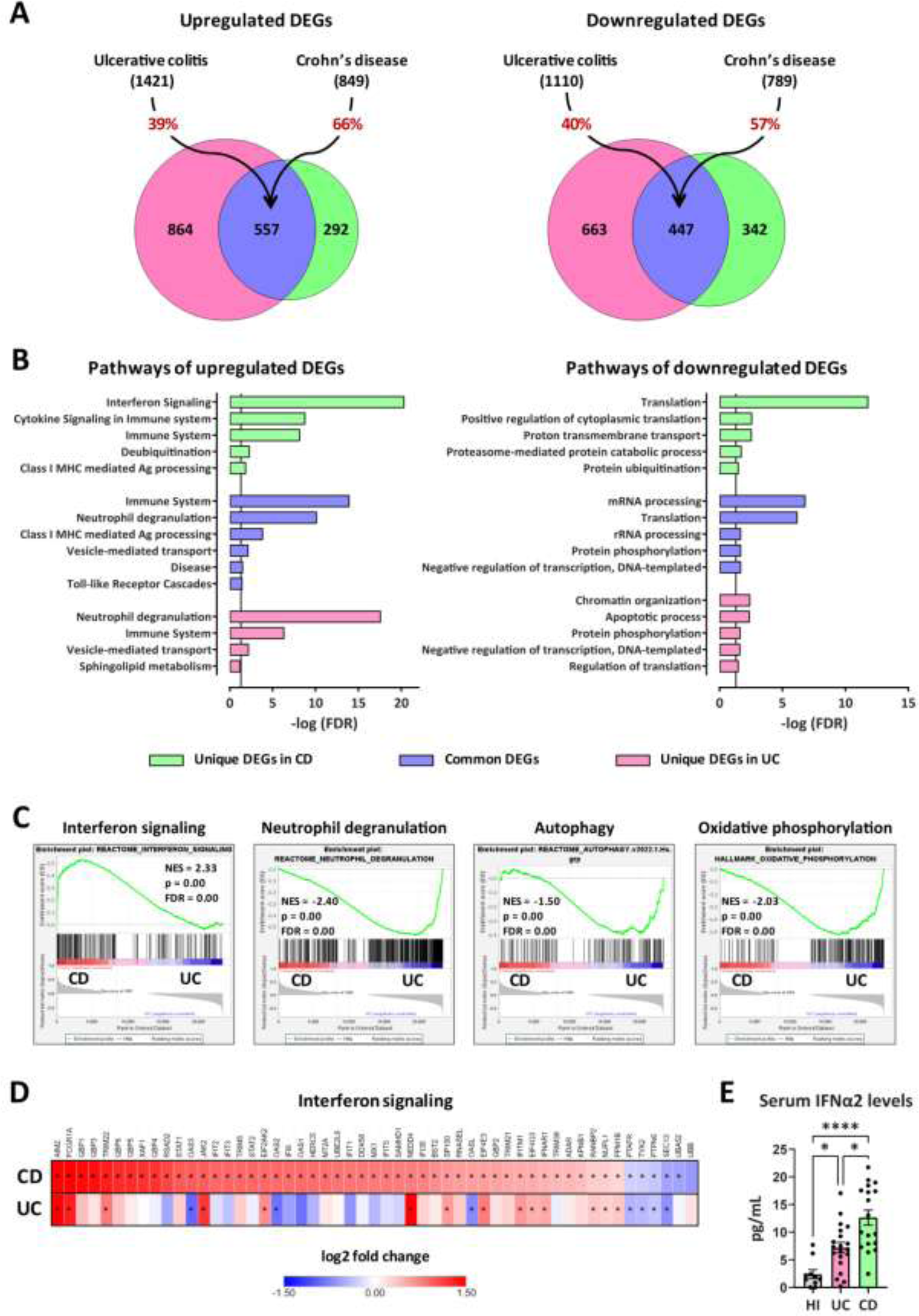
Transcriptomic analysis of peripheral blood neutrophils isolated from patients with CD or UC. **(A)** Venn diagrams of upregulated and downregulated DEGs (baseMean > 30 and FDR < 0.05), following RNA-Seq analysis of peripheral blood neutrophils isolated from patients with CD (*n=18*) or UC (*n=24*). DEGs were identified following comparison with neutrophils isolated from healthy individuals (*n=18*). The percentages on the arrows indicate the overlap between the two diseases. **(B)** Graphs depicting the top upregulated and downregulated pathways in neutrophils isolated from CD or UC patients. Reactome and GO Biological Process annotations were used for up- and down-regulated DEGs, respectively. Pathways with redundant sets of DEGs were excluded. Vertical lines show the threshold for statistical significance (FDR < 0.05). **(C)** GSEA plots of significantly altered signatures in the transcriptome of CD *versus* UC neutrophils, using Reactome and Hallmark as reference gene sets from the Human Molecular Signatures Database. **(D)** Heatmap depicting the log2 fold change values of Interferon signaling pathway (Reactome, R-HSA-913531.3), as determined by RNA-Seq analysis of CD and UC neutrophils. Asterisks depict statistical significance (FDR < 0.05). **(E)** Levels of IFNα2 in the serum of HI and patients with CD or UC. Data are expressed as Mean ± SEM. Nonparametric Kruskal-Wallis test was applied, followed by Dunn’s multiple comparisons test, *p < 0.05, and ****p < 0.0001. CD, Crohn’s disease; DEGs, differentially expressed genes; FDR, false discovery rate; GSEA, gene set enrichment analysis; HI, healthy individuals; NES, normalized enrichment score; RNA-Seq, RNA-Sequencing; UC, ulcerative colitis.

Gene set enrichment analysis (GSEA) was also performed, using the Reactome and Hallmark gene sets collections of the Human Molecular Signatures Database, to reveal enriched signatures in our datasets, and independently verify the overrepresentation of interferon signaling and neutrophil degranulation in CD and UC neutrophils, respectively (**Fig. 4C**). GSEA also highlighted the increased expression of DEGs involved in autophagy and oxidative phosphorylation in UC, compared to CD neutrophils, consistent with the previously reported increased NETotic potential in UC (16).

Focusing on the interferon signaling pathway, 53 DEGs were significantly regulated in CD neutrophils, including several target genes, intracellular mediators, enzymes, and receptors (**Fig. 4D**). Of those, 47 were up- and only six were down-regulated. On the other hand, only 14 out of 53 aforementioned genes were significantly upregulated in UC, while three major interferon-induced genes (*OAS2*, *OAS3*, and *OASL*) were downregulated (**Fig. 4D, Fig. S3E**). Moreover, RT-qPCR analysis in neutrophils from an independent group of CD and UC patients, confirmed that mRNA levels of the key IFN-signaling genes *STAT1* and *STAT2* followed the same expression pattern with transcriptome analysis (**Fig. S4A**). Hence, the molecular signature that distinguishes CD from UC neutrophils is positively correlated with interferon signaling activation.

Next, we sought to measure the levels of interferons, along with several other cytokines in the sera of patients with CD and UC. Although both IFNα2 and IFNγ were found elevated in the sera of IBD patients, in comparison to healthy individuals, only IFNα2 levels were significantly increased in CD over UC patients (**Fig. 4E, Fig. S4B**). Thus, elevated IFNα2 levels in the serum possibly account for the interferon fingerprint characterizing the transcriptome of peripheral neutrophils in CD.

### 2.5 IFNα primes neutrophils of CD patients to acquire fibrotic partnership

Based on the findings above, we hypothesized that IFNα may signal in neutrophils committing them to acquire a fibrotic partnership in their crosstalk with fibroblasts. To this end, we stimulated *in-vitro* HI neutrophils with serum from CD patients to produce eNETs (CD serum-generated eNETs), which were subsequently used for the treatment of HI primary fibroblasts.

We found that treated fibroblasts acquired a fibrotic phenotype; KLF2 (-), CCN2 (++), and collagen (++) (**Fig. 5A,B, I-K, Fig. S5A,B**), a phenomenon that was not observed when we used CD serum-generated eNETs obtained after neutralizing IFNα in serum (**Fig. 5C, I-K, Fig. S5A,B**), or when HI neutrophils were treated with the JAK-1/2 inhibitor baricitinib before their stimulation by CD serum to form eNETs (**Fig. 5E, I-K, Fig. S5A,B**). Neutralization of IFNα in the already formed CD serum-generated eNETs was ineffective in preventing fibroblasts from acquiring a fibrotic phenotype, suggesting the early, priming effect of IFNα on neutrophils (**Fig. 5D, I-K, Fig. S5A,B**).

**Figure 5.**
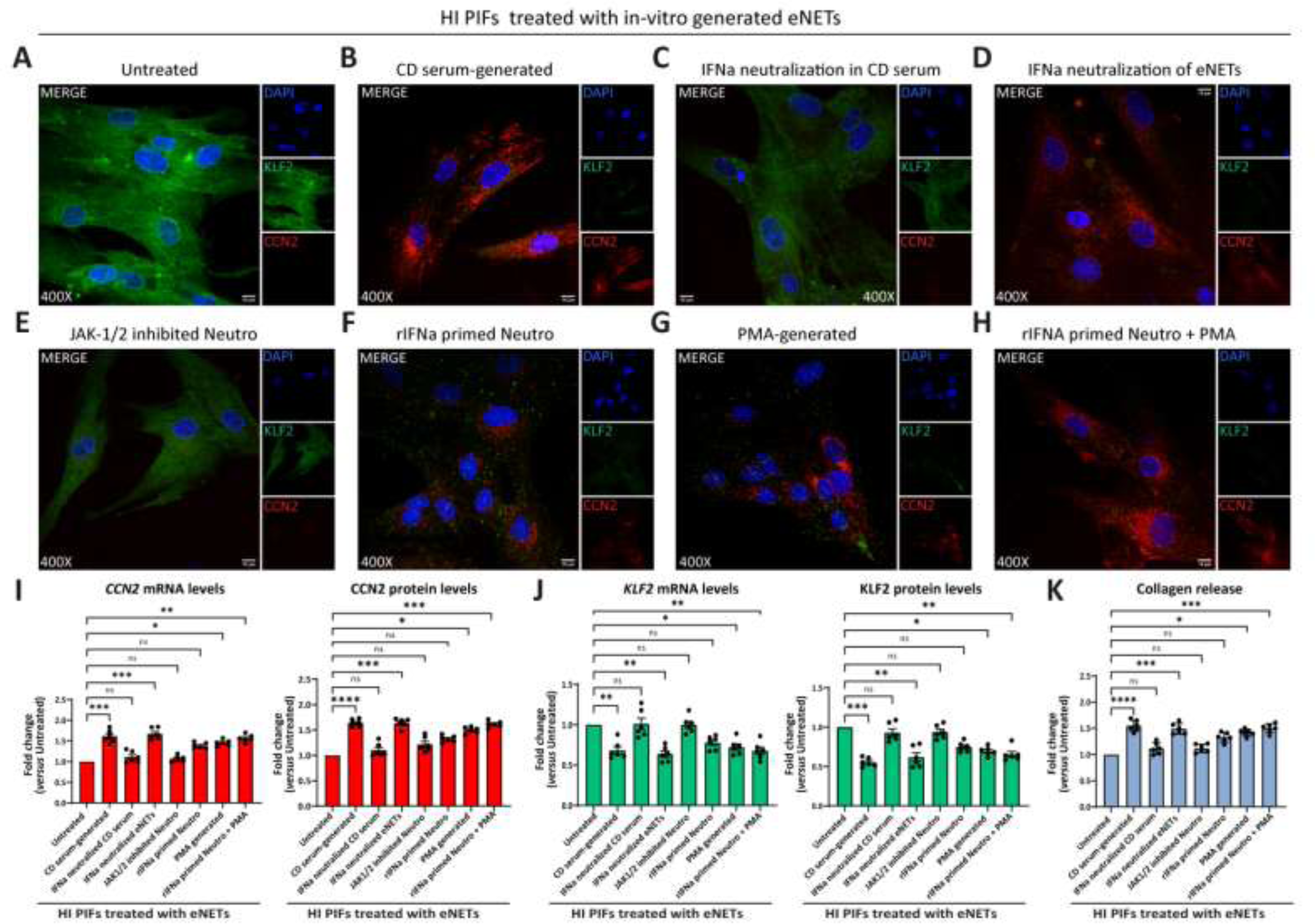
CD neutrophils are primed through IFNα/JAK signaling to exert their fibrotic role. **(A-H)** Evaluation of CCN2 and KLF2 by immunofluorescence (blue: DAPI, green: KLF2, red: CCN2) in HI PIFs treated with *in-vitro* generated eNETs as follows: **(A)** HI neutrophils stimulated by CD serum to form eNETs in the **(B)** absence or **(C)** presence of an IFNα neutralizing antibody. **(D)** IFNα neutralization of already formed CD-serum eNETs. **(E)** Inhibition of JAK-1/2 signaling in neutrophils with baricitinib. **(F)** Neutrophils primed with IFNα to produce eNETs. **(G)** PMA-generated eNETs. **(H)** Combination of **(F)** and **(G)** to produce eNETs that subsequently treated HI PIFs. **(I)** CCN2 and **(J)** KLF2 mRNA and protein levels of eNETs treated-fibroblasts, as described above, assessed by qPCR and in-cell ELISA. **(K)** The corresponding collagen release assay. **(A-H)** One representative example out of four independent experiments is shown. Confocal microscopy. Magnification: 400x, Scale Bar: 10μm **(I-K)** Nonparametric Kruskal-Wallis followed by Dunn’s multiple comparisons test, *n=6*, *p<0,05, **p<0,01, ***p<0,001, ****p<0,0001, n.s.: not significant. Data are expressed as mean ± SEM. CCN2, cellular communication network factor 2; CD, Crohn’s disease; eNETs, enriched neutrophil extracellular traps; HI, healthy individuals; IFNa, Interferon alpha; JAK, Janus kinase; KLF2, Kruppel-like factor 2; MFI, mean fluorescence intensity; PIFs, primary intestinal fibroblasts; PMA, Phorbol-12-myristate-13-acetate.

Furthermore, healthy fibroblasts that had acquired a KLF2 (+), CCN2 (+), collagen (+) mild fibrotic phenotype upon treatment with PMA-generated eNETs, were transformed towards KLF2 (-), CCN2 (++), collagen (++) fibrotic active cells, when recombinant IFNα primed the neutrophils during the procedure of PMA eNETs generation (**Fig. 5F-K, Fig. S5A,B**).

Collectively, our data so far suggest that in CD, priming of circulating neutrophils by IFNα is essential to commit their fibrotic effect over intestinal fibroblasts and their plasticity is expressed through eNETs.

### 2.6 Neutrophil-dependent fibrosis in CD is eliminated by dismantling the DNA scaffold of eNETs or by inhibiting JAK signaling in fibroblasts

Prompted by the above-mentioned findings indicating that IFNα primes neutrophils to acquire a fibrotic role through eNETs and since previous results showed that JAK/signaling in fibroblasts is involved in COVID-19 immunofibrosis (13), we tried to diminish the fibrotic transformation of fibroblasts either by dismantling the DNA scaffold of eNETs or by inhibiting JAK signaling in these cells.

Treatment of CD eNETs with DNaseI or pretreatment of HI fibroblasts with baricitinib inhibited the transformation of the KLF2 (++), CCN2 (-), collagen (-) HI fibroblasts towards the KLF2 (-), CCN2 (++), collagen (++) CD-like phenotype (**Fig. 6A-D, F-I**). This effect was also observed with the plant-derived polyphenol tannic acid, a potent inducer of KLF2 expression (13, 17), suggesting a potential role of KLF2 in the fibrotic process (**Fig. 6E-I).** Next, we investigated whether JAK inhibition could reverse to normal the fibrotic phenotype of the already affected primary fibroblasts of CD. The KLF2 (-), CCN2 (++), collagen (++) phenotype of active CD primary fibroblasts was not altered following their treatment with baricitinib, suggesting that JAK inhibition should be performed early, before acquiring the CD phenotype (**Fig. S6A-E**).

**Figure 6.**
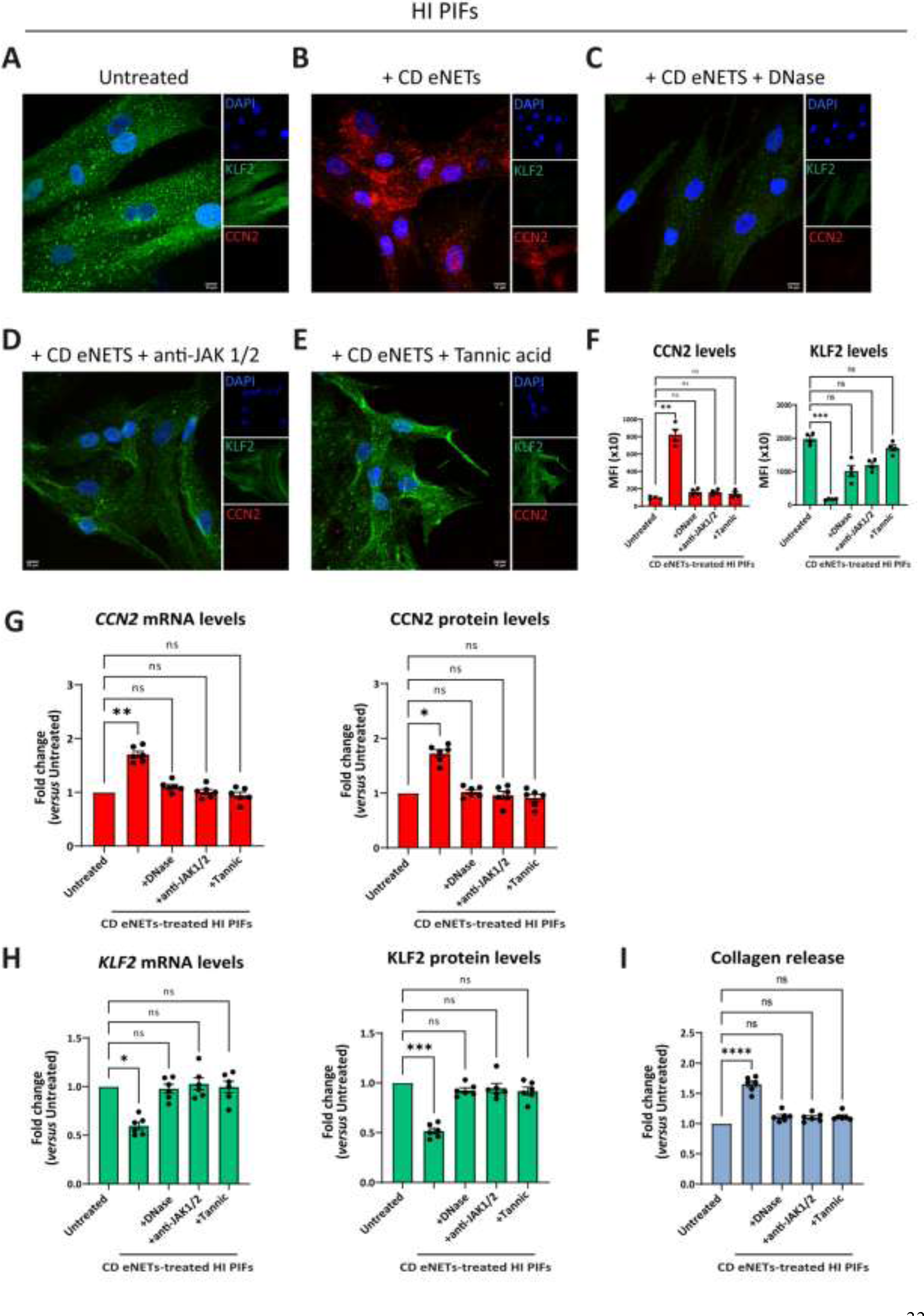
Disruption of NET-scaffold or inhibition of JAK-1/2 signaling on fibroblasts prevents neutrophil-mediated fibrosis. **(A-E)** Assessment of CCN2 and KLF2 by immunofluorescence (blue: DAPI, green: KLF2, red: CCN2) and **(F)** MFI quantification, in stimulation and inhibition studies. **(A)** HI PIFs, treated with **(B)** *ex-vivo* CD eNETs or **(C)** CD eNETs pre-treated of with DNaseI to dismantle the DNA-scaffold. **(D)** Pre-treatment of HI PIFs with JAK-1/2 inhibitor baricitinib and subsequent stimulation with CD eNETs. **(E)** Tannic acid, a chemical inducer of KLF2, was used as a positive control of KLF2 expression. **(G)** Analysis of CCN2 and **(H)** KLF2 mRNA and protein expression, and **(I)** Collagen release assay, in PIFs of the above mentioned *in-vitro* studies. **(A-E)** One representative example out of four independent experiments is shown. Confocal microscopy. Magnification: 400x, Scale Bar: 10μm. **(F-I)** Nonparametric Kruskal-Wallis followed by Dunn’s multiple comparisons test, *n=6*, *p<0,05, **p<0,01, ***p<0,001, ****p<0,0001, n.s.: not significant. Data are expressed as mean ± SEM. CCN2, cellular communication network factor 2; CD, Crohn’s disease; eNETs, enriched neutrophil extracellular traps; ΗΙ, healthy individual; JAK, Janus kinase; KLF2, Kruppel-like factor 2; MFI, mean fluorescence intensity; STAT, signal transducer and activator of transcription.

In conclusion, neutrophil-fibroblast crosstalk in CD is disrupted by targeting neutrophil DNA scaffold or/and JAK signaling in fibroblasts before their interaction. This neutrophil-driven fibrotic process seems to be an early event in the pathogenesis of CD immunofibrosis.

### 2.7 Levels of interferon-signaling components are positively correlated with the severity of CD

To further support our findings, we sought to examine whether serum IFNα levels and transcriptomic alterations of IFN-signaling-related genes in peripheral neutrophils are associated with the severity of CD, as reflected by the classical disease activity index, CDAI (18).

We observed that IFNα2 levels in the serum of CD patients were correlated with the disease severity, since individuals with higher CDAI scores were also characterized by higher concentrations of IFNα2 (**Fig. 7A**). In contrast to CD, in UC no correlation was observed between IFNα levels and Mayo DAI score, a common indicator of disease activity (19) (**Fig. 7A**).

**Figure 7.**
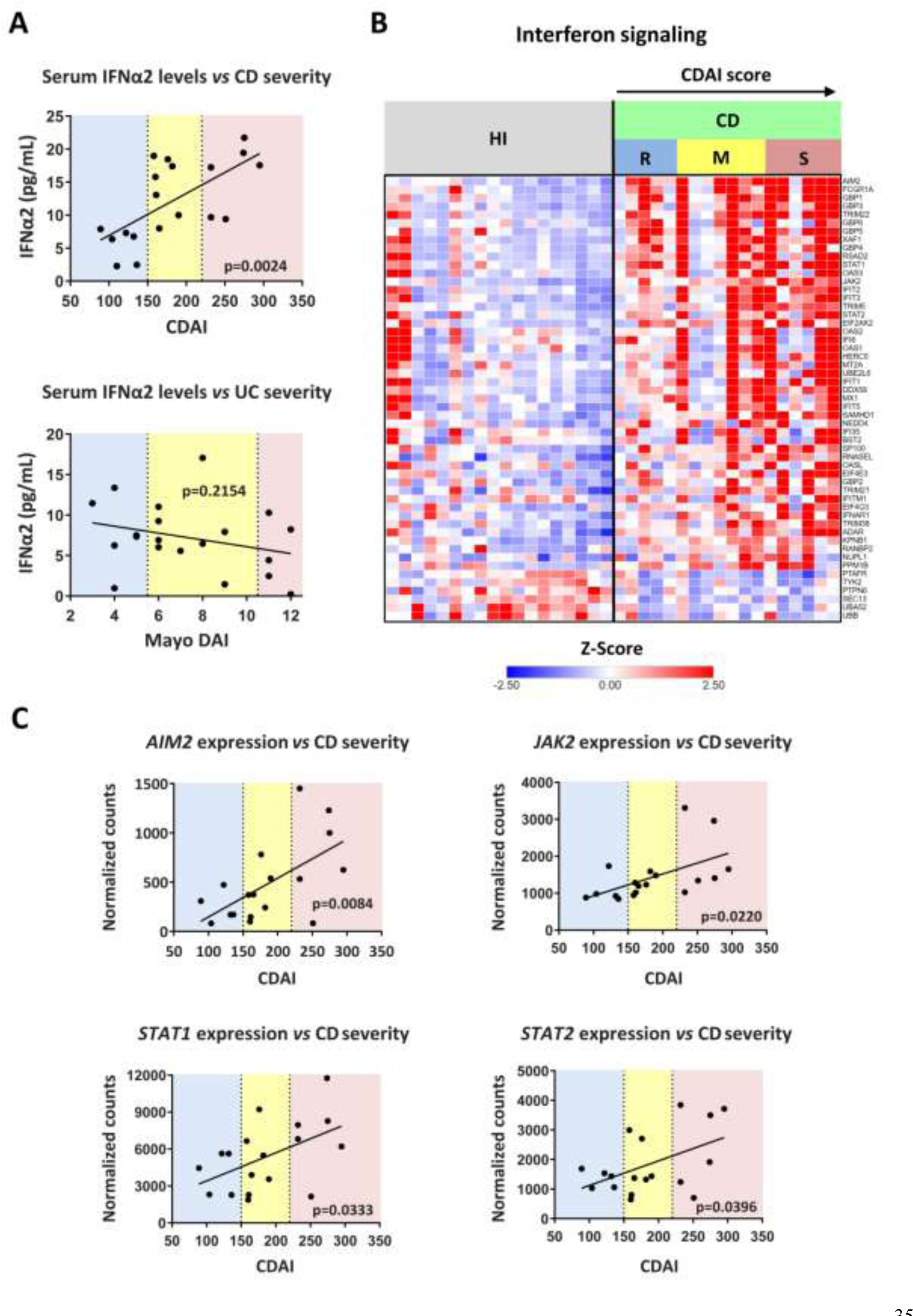
Levels of interferon signaling components are positively correlated with Crohn’s disease severity. **(A)** Correlation plots of serum IFNα2 levels *versus* disease severity in CD (upper graph) and UC (lower graph). CDAI and Mayo DAI were used for assessing disease activity in CD and UC, respectively. **(B)** Heatmap depicting the relative expression of DEGs belonging to Interferon signaling pathway (Reactome, R-HSA-913531.3), as determined by RNA-Seq analysis of neutrophils isolated from HI (*n=18*) and CD patients (*n=18*). **(C)** Correlation plots of the mRNA expression of key interferon signaling components (*AIM2*, *JAK2*, *STAT1*, and *STAT2*), as determined by RNA-Seq analysis of CD neutrophils, *versus* disease severity in CD. Simple linear regression was used in all panels to assess the relationship between the studied variables. CD, Crohn’s disease; CDAI, Crohn’s disease activity index (R, remission, < 150; M, mild to moderate, 150-220; S, moderate to severe, > 220); HI, healthy individuals; RNA-Seq, RNA-Sequencing; UC, ulcerative colitis.

We also noticed that the transcriptomic alterations observed in peripheral blood neutrophils of IBD patients were also correlated with the CDAI and Mayo DAI scores (**Fig. S7A,B**). The whole set of DEGs followed an expression pattern related to the disease severity (**Fig. S7A,B**), an observation that was prominent for interferon signaling components in CD as well (**Fig. 7B**). Indeed, the mRNA expression of key components of the interferon signaling pathway, namely *AIM2*, *JAK2*, *STAT1*, and *STAT2*, followed faithfully the CDAI score since it was increased relative to disease severity (**Fig. 7C**). The expression of several interferon-signaling components was also correlated with serum IFNα2 levels (**Fig. S7C**).

Taken together, serum IFNα and/or mRNA levels of interferon-signaling components in neutrophils emerge as potential diagnostic/prognostic biomarkers for CD.

## 3. DISCUSSION

In this study, we describe a novel mechanistic link between neutrophils and fibroblasts that characterizes immunofibrosis of CD. In contrast to UC, neutrophils of CD patients, exhibit pro-fibrotic properties by activating intestinal fibroblasts leading to increased collagen release. This type of neutrophil plasticity is primed by IFNα signaling and expressed by the NET-enriched inflammatory environment. Moreover, neutrophils may be attracted to the intestinal environment by IL-8 secreted from activated fibroblasts sustaining this fibrotic loop of neutrophil-fibroblast interaction. Further supporting this mechanistic basis, the expression levels of key IFNα pathway components in serum and neutrophils are well correlated with the clinical activity of CD patients.

Growing evidence today implies that the development of tissue fibrosis involves the complex interplay between immune and stromal cells. However, in CD, cell-cell interactions and functions are incompletely understood, while most of the studies focus mainly on lymphocytes and/or other mononuclear immune cells, while the role of neutrophils remains obscure (14).

Here, we showed that neutrophils in active CD and UC may be recruited at the site of tissue damage by IL-8 produced by intestinal fibroblasts. Earlier clinical studies have indicated increased IL-8 levels in the intestinal mucosa of active IBD patients (20, 21). This study additionally characterizes fibroblasts as a source of IL-8 in the tissue environment of IBDs suggesting a functional, chemoattractant, effect on peripheral neutrophils. Future studies should aim to elucidate the specific mediator(s) responsible for initiating IL-8 expression in fibroblasts, as well as explore the putative role of gut microbiota dysbiosis. Similar triggers might also induce IL-8 production from connective tissue mesenchymal cells in IBD extraintestinal manifestations, such as arthritis. In support of this, it has been shown previously, that NETs from rheumatoid arthritis patients can stimulate the production of IL-8 from fibroblast-like synoviocytes (10). Of note, although fibroblast-derived IL-8 appears to be implicated in neutrophil recruitment in both IBDs, we observed a differential intestinal distribution of neutrophils. In CD, dense neutrophil infiltrations were observed in proximity to fibrotic areas, implying a fibrotic role for recruited neutrophils. This finding combined with the differences in the plasticity of neutrophils between the two IBDs, could explain the fibrotic phenotype of CD.

Currently, the traditional concept that neutrophils comprise terminally differentiated cells with limited plasticity and highly conserved function has been critically revised (5, 6). Several studies support that during inflammatory conditions, the transition of mature neutrophils from bone marrow to the bloodstream is accompanied by changes at the transcriptional level that enable the acquisition of distinct functions within the affected tissues (5, 6). Previous clinical and experimental studies suggested that in the context of different inflammatory disorders, neutrophils may acquire differential plasticity which may be reflected by the NETs that they release (9, 10). In this context, our group and others have indicated that activated neutrophils may acquire an immunofibrotic role through the release of NETs. These fibrogenic NETs were able to activate human fibroblasts, inducing their proliferation, differentiation to myofibroblasts and immunogenicity (10–12).

Our functional studies indicated that NET-enriched extracellular mediators (eNETs) *ex-vivo* isolated from peripheral neutrophils of CD patients, and not the CD serum directly, were able to transform healthy intestinal fibroblasts toward the distinct CD phenotype characterized by negative KLF2 and high CCN2 expression, leading to collagen production. A similar immunofibrotic phenotype of lung fibroblasts has been also described in severe COVID-19 (13). However, eNETs from peripheral neutrophils of UC patients did not have a similar potential, suggesting differential neutrophil/NETs plasticity on fibroblasts between the two IBDs.

To identify possible molecular pathways and targets that may drive this plasticity, we applied whole transcriptome analysis in peripheral blood neutrophils from IBD patients. Importantly, although a substantial overlap of the DEGs between the two intestinal diseases was observed, peripheral neutrophils of CD were selectively characterized by an IFN-responsive signature. On the other hand, in UC, upregulated genes were enriched for pathways related to neutrophil degranulation, autophagy and oxidative phosphorylation, confirming our previous mechanistic studies which demonstrated the key role of autophagy-mediated NET formation in UC (16). Recently, top-upregulated severity genes in the colonic mucosa of UC patients have been also found to be involved in innate immunity and neutrophil degranulation (22). Previous comparative transcriptome studies between UC and CD have yielded heterogeneous results mostly being non-targeted, performed in whole blood or mucosal tissue (23–25). Despite this, a mostly neutrophil-like signature has been proposed for the whole blood of IBD patients, while the most significant signal within CD ileal mucosa with deep ulcers was for granulocytes, further favoring the role of neutrophils in active IBD (23, 25). At the time of this writing, emerging single-cell transcriptome data in mucosal samples reveal the heterogeneity of tissue neutrophils in IBD patients. Noteworthy, A. Garrido-Trigo *et al.* report that, in contrast to UC, the majority of intestinal neutrophils in CD display a marked IFN-response signature (26). This is in agreement with, and further supports, our transcriptome findings in peripheral neutrophils of CD.

Based on the findings indicating that IFN signature may distinguish CD from UC neutrophils, and multiplex cytokine analysis showing that IFNα, and not IFNγ, serum levels were significantly increased in CD compared to UC, it was reasonable to assume that IFNα may prime peripheral neutrophils of CD patients to exert their fibrotic effect on fibroblasts. In line with this, functional studies indicated that CD serum induces the generation of highly fibrogenic eNETs in an IFNα-dependent manner. Furthermore, neutrophils primed with IFNα, during the production of PMA-generated eNETs, enhanced their fibrotic plasticity. This further supports an additional key role for IFNα in the immunofibrotic plasticity of neutrophils, which is expressed *via* extracellular DNA structures. A large body of evidence has demonstrated that the integrity of DNA scaffold is important for NETs function (10, 11, 27). How the architecture of NETs and their proteins are implicated in the immunofibrotic plasticity of neutrophils is an intriguing question that needs to be subjected to further investigation in the future.

Several studies have indicated that loss-of-function mutations in nucleotide-binding oligomerization domain 2 (NOD2) have been associated with CD (28). Of interest, it has been recently indicated, that NOD2 activation in hematopoietic cells protected mice from TLR9-induced exacerbation of DSS-induced colitis by downregulating IFNα responses (29). Although several immune and non-immune cells may contribute, the exact sources of IFNα in the systemic circulation of CD remain to be identified. It has been previously shown that mature neutrophils in systemic lupus erythematosus (SLE) are primed *in-vivo* by type I IFNs to release NETs upon exposure to SLE-derived autoantibodies, indicating plasmacytoid dendritic cells as a major source of IFNα (30).

Fibrosis leads to abnormal tissue remodeling complicating several chronic inflammatory diseases, while early prevention of fibrosis remains a significant unmet medical need (31). Our mechanistic studies suggest that the interplay between neutrophils/eNETs and fibroblasts in CD is an early immunofibrotic event, that may be disrupted by using various pharmaceutical agents, such as anti-IL-8 (32) to interrupt the intestinal migration of pro-fibrotic neutrophils, recombinant DNaseI to dismantle the chromatin scaffold of fibrogenic eNETs (33) or inhibitors of JAK signaling (34) both in peripheral neutrophils and intestinal fibroblasts to prevent their activation (**Fig 8**). Recent experience in severe COVID-19 patients indicates that combining different immunomodulatory therapies may be beneficial for complex fibrotic diseases such as CD (13, 33). Lately, JAK inhibition has been approved as a new treatment for moderate-to-severe CD patients, further supporting the translational impact of our study (34).

In several cases, the assessment of disease activity or the discrimination between UC and CD is problematic, mainly based on invasive interventions such as ileocolonoscopy and intestinal biopsies (1). Linking our findings with clinical practice, we found that levels of IFNα in serum or/and mRNA expression of selective IFN-signaling-related components in peripheral neutrophils could be surrogate markers of CD activity, positively correlated with the CDAI, a well-established disease severity index which includes endoscopic findings. Recently, colonic tissue biopsies of CD or UC patients with active and remitted disease phases were characterized by the enhanced and reduced expression of IFN-stimulated genes, respectively (29). Here, we suggest that targeted analysis of peripheral blood neutrophils may be a more specific, easier, and less costly diagnostic approach.

In conclusion, this study unravels the role of IFNα/JAK signaling in the plasticity of neutrophils/NETs during their crosstalk with intestinal fibroblasts, eventually leading to immunofibrosis of CD. The IFNα/neutrophil/fibroblast pathway provides novel candidate targets for the design of future diagnostic and therapeutic strategies in CD.

## 4. MATERIALS AND METHODS

### 4.1 Patients and sampling

The study was conducted in the First Department of Internal Medicine and the Gastroenterology-Hepatology Unit, at the University Hospital of Alexandroupolis. In total 26 treatment-naïve CD (16 male/10 female; mean age 35.3±15.0 years), 32 UC patients (24 male/8 female; mean age 52.2±19.1 years) and 18 healthy individuals including 4 who underwent preventive screening colonoscopy (HI; 12 male/6 female; mean age 39.1±12.6 years), were recruited. The diagnosis of UC and CD was according to standard clinical, endoscopic, radiological, and histological criteria (1, 35). Clinical severity and disease behavior scores such as Mayo disease activity index (Mayo-DAI) (19), Crohn’s disease activity index (CDAI) (18) and Montreal score (36) were evaluated in both UC and CD patients by two independent, expert gastroenterologists.

Neutrophils, serum, primary intestinal fibroblasts (PIFs) and intestinal biopsies were collected. The baseline clinical characteristics of the patients and samples used in experimental procedures are depicted in **Supplementary Tables S1-3.**

### 4.2 Isolation of peripheral blood neutrophils and serum

Peripheral heparinized blood and serum were collected as previously described (16). Detailed methods are included in the Supplementary Materials and Methods.

### 4.3 Isolation, culture, and characterization of human primary intestinal fibroblasts

Detailed methods are included in the Supplementary Materials and Methods.

### 4.4 Generation and collection of enriched-neutrophil extracellular traps (eNETs)

A total of 1.5 × 10^6^ neutrophils isolated from UC, CD patients and HI were re-suspended in Roswell Park Memorial Institute (RPMI) medium (21875; Thermo Fisher Scientific; Carlsbad, SA, USA) supplemented with 2% heat-inactivated fetal bovine serum (FBS);10082147; Thermo Fischer Scientific), and cultured at 5% CO_2_, at 37°C, for 3.5 hours, based on the standard isolation protocol to generate *ex-vivo* NET structures. Similarly, healthy neutrophils were also incubated *in-vitro* in the presence of 5% serum from UC, CD patients and HI, or phorbol 12-myristate 13-acetate (PMA) (40 ng/ml; Sigma-Aldrich, St Louis, MO, USA), a generic inducer of NET release, and cultured in the aforementioned conditions. After the incubation period we performed a vigorous agitation for 5 minutes, to detach NET structures and this medium was collected. We omit the washing step, which was carried out during the classic method of NETs isolation (37–39) in order to collect more inflammatory mediators than NETs (10, 40, 41). This mixture of enriched NETs is hereafter referred to as “eNETs”. Aliquots of eNETs were stored at −80°C until analyzed. Both *ex-vivo* and *in-vitro* generated eNETs were further used in stimulation studies in PIFs at a final concentration of 20%.

To quantify NETs, MPO/DNA complexes were measured in *ex-vivo* and *in-vitro* NETs isolated from 1.5 x 10^6^ neutrophils as previously described. In brief, NETs were captured by human anti-MPO antibody (1:500 dilution; HM2164; clone 6G3-mouse IgG1; Hycult Biotech; Uden, Netherlands), and an anti-double-stranded DNA antibody was used for DNA detection (Cell Death Detection ELISA Kit; 11544675001; Merck; Kenilworth, New Jersey, USA). Absorbance was measured at 405 nm (42, 43).

The concentrations and time points used to test neutrophil function were optimized before the experiments. All materials used were endotoxin-free, as determined by a Limulus amebocyte assay (E8029; Sigma-Aldrich).

### 4.5 Stimulation and inhibition studies in cultured cells

PIFs or peripheral blood neutrophils were seeded into 6-well culture plates (≈0.8-1 x 10^6^ cells/well for fibroblasts, ≈1.5 x 10^6^ cells/well for neutrophils; Corning Incorporated) in complete DMEM and RPMI medium respectively.

Neutrophils were stimulated with recombinant IFNα2 (1000 U/mL; H6041; Sigma-Aldrich), in order to prime them to release eNETs. PIFs were stimulated with 5% serum from HI, UC, and CD patients, in complete DMEM. To block the IFN signaling, a neutralizing mouse monoclonal antibody against human Interferon Alpha (anti-IFNa; 15 μg/mL; 21100-1; R&D Systems;) was used in both neutrophils and PIFs. To inhibit Janus kinase signaling (JAK-1 and JAK-2), PIFs and neutrophils were pre-treated (60 min) with Baricitinib (2.5 nM; 16707; Cayman Chemical). To dismantle NET structures, CD *ex-vivo* eNETs were pre-incubated (60 min) with DNaseI (1 U/ mL; EN0525, Thermo Fisher Scientific). To trigger the expression of KLF2, PIFs were further pre-incubated (60 min) with Tannic acid (20 nM; 403040; Sigma-Aldrich), a polyphenolic compound, acting as a potent inducer of KLF2. The total culture period of neutrophils was 3.5 hours.

### 4.6 RNA isolation, cDNA synthesis and RT-qPCR

Detailed methods are included in the Supplementary Materials and Methods.

### 4.7 RNA sequencing and bioinformatics analysis

One μg of total RNA was used for the preparation of cDNA libraries, as previously described (44, 45). Sequencing was performed in a single-end manner at the Genome Center of Biomedical Research Foundation Academy of Athens, using the NovaSeq 6000 SP 100c kit (Illumina, 20028401), generating 100 bp long reads.

Raw sequence data were uploaded to the Galaxy web platform (46), and standard tools of the public server “usegalaxy.org” were used for subsequent analysis, as previously described (45). HISAT2 (v2.2.1+galaxy1) was applied for the alignment of trimmed reads to the human GRCh37/hg19 genome assembly from the Genome Reference Consortium, using the default parameters. Assessment of uniform read coverage for exclusion of 5’/3’ bias and evaluation of RNA integrity at the transcript level were performed, as previously reported (45). Moreover, absolute de-convolution of human immune cell types was applied in our datasets, according to the Shiny app, https://giannimonaco.shinyapps.io/ABIS (47), to ensure their enrichment in granulocytes gene expression signature (> 95%). Replicates with signatures enriched in contaminating lymphocytes (> 5%) were excluded from the analysis. The identification of Differentially Expressed Genes (DEGs) between HI and patients with either CD or UC, was carried out with the DESeq2 algorithm (v2.11.40.7+galaxy2) (48), using the count tables generated from htseq-count tool (v0.9.1+galaxy1) as input.

Pathway analysis was performed using the GeneCodis4 web-based tool (49). Cutoff values for statistically significant DEGs were baseMean >30 and adjusted p-value (false discovery rate, FDR) <0.05. Gene set enrichment analysis (GSEA) was performed using the GSEA software (University of California, San Diego & Broad Institute, USA) (50). Briefly, normalized counts generated from DESeq2 algorithm and annotated gene sets from the Human Molecular Signatures Database (Human MSigDB v2023.1) were used as inputs. Gene sets were ranked by taking the -log10 (p-value) and signed as positive or negative based on the direction of fold change, followed by pre-ranked analysis using the default settings (1000 permutations, min, and max term size of 15 and 500, respectively).

Heatmaps were generated using the Morpheus software, https://software.broadinstitute.org/morpheus (Broad Institute, USA). Venn diagrams were constructed with Venny 2.1 (developed by Oliveros, J.C., 2007) and Venn Diagram Plotter software (Pacific Northwest National Laboratory, U.S. Department of Energy).

### 4.8 In-cell ELISA (ICE assay, Cytoblot)

Details are included in the Supplementary Materials and Methods.

### 4.9 Immunofluorescence staining in human PIFs

Details are included in the Supplementary Materials and Methods.

### 4.10 Collagen measurement

Details are included in the Supplementary Materials and Methods.

### 4.11 Multiplex cytokine measurement

Details are included in the Supplementary Materials and Methods.

### 4.12 Immunohistochemistry (IHC-P), Masson’s trichrome and Immunofluorescence (IF) staining in tissue sections

Detailed methods are included in the Supplementary Materials and Methods.

### 4.13 *In-vitro* transwell migration assay (chemotaxis assay)

Details are included in the Supplementary Materials and Methods.

### 4.14 Statistical analysis

Statistical analysis was performed with the GraphPad Prism software (version 9.0, San Diego, CA, USA). To analyze more than two groups, the nonparametric Kruskal-Wallis test, followed by Dunn’s test, was performed. To compare the migratory capacity of HI PIFs supernatants to UC and CD, Bayesian unpaired t-test was used; p-values were adjusted using the Benjamini-Hochberg correction. For comparisons between IL-8 neutralized PIFs supernatants and untreated supernatants, Bayesian paired t-tests were performed, using the JZS Bayes factor (**Fig. 2G**) (51). Data are expressed as the mean ± standard error of the mean (SEM). Simple linear regression was used to assess the relationship between two variables. The level of significance was set to *p<0.05, **p<0.01, ***p<0.001, ****p<0.0001.

### 4.15 Study approval

All study participants provided written informed consent in accordance with the principles expressed in the Declaration of Helsinki. Patients’ records were anonymized and de-identified prior to analysis to ensure anonymity and confidentiality. The study protocol was approved by the Scientific and Ethics Committee of the University Hospital of Alexandroupolis (Approval Number 803/23-09-2019).

### 4.16 Data availability

Raw data supporting the findings of this study are available from the corresponding authors on request. The datasets generated for this study can be found in the NCBI Sequence Read Archive (https://www.ncbi.nlm.nih.gov/sra), accession number PRJNA997815.

## Author contributions

Conceptualization: K.R., P.S., Methodology: E.G., G.D., AM.N., V.P., Investigation: E.G., G.D., AM.N., N.K., D.K., C.A., E.S., E.P., M.A.K., V.T., AR.G., M.A., AL.G., G.K., Visualization: E.G., G.D., AM.N., S.D., AL.G., Funding acquisition: M.K., P.S., Project administration: P.S., Supervision: K.R., P.S., Writing – original draft: E.G., G.D., AM.N., K.R., P.S., Writing – review & editing: E.G., G.D., AM.N., N.K., C.A., M.A.K., M.A., D.T., M.K., G.K., K.R., P.S.

## Acknowledgments

This study was supported by the Greek General Secretariat for Research and Innovation (GSRI), Research & Innovation Programme CytoNET, grant MIS-5048548 and by GSRI, Regional Excellence Programme InTechThrace, grant MIS-5047285.

We gratefully acknowledge Dr. P. Sideras for his support and advices during this project, and Dr. I. Theodorou and Dr. GS. Krasnov for their suggestions regarding RNA-seq analysis. We kindly thank Ms. Kyriaki Devetzi for her technical support. Graphical abstract was created with Biorender.com.

## SUPPLEMENTAL MATERIAL

### Supplemental Materials and Methods

### Isolation of peripheral blood neutrophils and serum

Peripheral heparinized blood was collected and circulating neutrophils were isolated by Histopaque double-gradient density centrifugation (1.119,1 g/mL and 1.077,1 g/mL, Sigma-Aldrich, St Louis, MO, USA) at 700 ×g, for 35 min at 20-25°C. Then, cells were washed once with phosphate-buffered saline (PBS-1×, Biosera) and centrifuged at 200 ×g for 12 min before being cultured. Neutrophils were adjusted to the desired concentration and used within 2 hours after isolation. Cell purity was estimated ≥98%.

To collect serum, venous blood from either HI or patients with CD and/or UC was collected in appropriate BD Vacutainer® Plus Plastic Serum tubes (Becton, Dickinson, and Company) and centrifuged at 500 ×g for 15 min. Serum samples were stored at −80°C until analyzed.

### Isolation, culture, and characterization of human primary intestinal fibroblasts

To investigate the pro-fibrotic potential of the intestinal inflammatory environment, human primary intestinal fibroblasts (PIFs) were isolated from colonic and/or terminal ileum biopsies obtained by colonoscopy from inflammatory lesions of active UC and CD patients as described above, based on a standard lab protocol (52, 53). PIFs isolated from healthy subjects who underwent preventive colonoscopy, served as controls. PIFs in passages 3–6 were cultured in DMEM (31885-023; Thermo Fisher Scientific) supplemented with 10% heat-inactivated fetal bovine serum (FBS; 10082147; Thermo Fischer Scientific), 200 U/mL antibiotic/antimycotic solution (15240062; Thermo Fisher Scientific), 1% non-essential amino acids (11140-035; Thermo Fisher Scientific) at 37 °C, 5% CO_2_ (complete DMEM). To verify their phenotype, cells were stained using antibodies against alpha-smooth muscle actin (α-SMA), vimentin, and desmin (54). PIFs were also plated in 6-well culture plates (≈0.8-1 x 10^6^ cells/well; Corning Incorporated) in complete DMEM to collect their supernatant. Once cells were 60% to 70% confluent, the medium was removed, cells were rinsed in PBS, and fresh low-serum DMEM (2% FBS) medium was added for 24 hours. After 24 hours, the medium was collected, centrifuged and the supernatant was transferred to a new tube. Aliquots of the supernatant were stored at −80°C until use.

### RNA isolation, cDNA synthesis and RT-qPCR

RNA was extracted from PIFs and peripheral blood neutrophils in TRIzol reagent following the manufacturer’s protocol (15596026; Thermo Fischer Scientific). Equal amounts of RNA were used to synthesize cDNA for Real-time qPCR (RT-qPCR) analysis or to prepare cDNA libraries for RNA-Sequencing. cDNA for RT-qPCR was generated using PrimeScript^TM^ RT Reagent Kit (RR037A; Takara) according to the manufacturer’s instructions. RT-qPCR for Krüppel-like Factor 2 (*KLF2*), Interleukin-8 (*IL8*), Signal transducer and activator of transcription 1 (*STAT1*), Signal transducer and activator of transcription 2 (*STAT2*) or/and Cellular Communication Network Factor 2 (*CCN2*) was performed by a KAPA SYBR FAST qPCR Master Mix (2X) (KK4602; KAPA Biosystems). To normalize the expression of the above-mentioned genes, glyceraldehyde 3-phosphate dehydrogenase (*GAPDH*) was used as an internal control gene. Details regarding the primer pairs and conditions of RT-qPCR are given in **Supplementary Table S4**. The data were quantified and analyzed using the 2^-ΔΔCt^ mathematical model.

### In-cell ELISA (ICE assay, Cytoblot)

To study the intracellular protein levels of CCN2 or KLF2, PIFs were cultured in 96-well high-binding microplates (655061; Thermo Fisher Scientific) in the presence of distinct stimuli for 4 h, as previously described (13, 37, 55). Briefly, goat anti-CCN2 (2 μg/mL; sc14939; Santa Cruz Biotechnology Inc) or mouse anti-KLF2 (6 μg/mL; MA5-24300; Thermo Fisher Scientific) were used as primary monoclonal antibodies. Horseradish peroxidase-conjugated donkey anti–goat IgG (1:1000 dilution, HAF109; R&D Systems) or donkey anti–mouse IgG (1:1000 dilution, HAF018; R&D Systems) were used as secondary antibodies. Absorbance was measured at 620 nm. The corrections were performed by subtracting the signal of the wells incubated in the absence of the primary antibody.

### Immunofluorescence staining in human PIFs

PIFs were cultured in 8-well-chambered slides (80806; Ibidi), and treated with various stimuli for 4 h, as previously described (13, 33). In brief, a rabbit anti-CCN2 polyclonal antibody (1:600 dilution; ab5097; Αbcam), a mouse anti-KLF2 monoclonal antibody (6 μg/mL; MA5-24300; Thermo Fisher Scientific), a mouse anti-IL-8 monoclonal antibody (15 μg/mL; MAB208-100; R&D systems), a rabbit anti-desmin polyclonal antibody (1:200; ab15200; Abcam), and/or a rabbit anti-Vimentin monoclonal antibody (1:250; ab92547; Abcam), were used as primary antibodies. Afterward, a Drop-n-Stain CF488A Goat anti-mouse IgG (H+L), highly cross-absorbed antibody (20956; Biotium) or a Drop-n-Stain CF594 Goat anti-rabbit IgG (H+L), highly cross-absorbed antibody (20955; Biotium) were utilized as the secondary antibodies. A mounting medium with DAPI (50011; Ibidi) was used in the final step. To distinguish specific staining from non-specific binding or background fluorescence, no primary antibody staining was used as internal control in each experiment.

Imaging was performed on a customized Andor Revolution Spinning Disc Confocal system with Yokogawa CSU-X1 (Yokogawa, Tokyo, Japan) confocal scan-head, build around an Olympus IX81 microscope (Olympus, Tokyo, Japan) mounted with a 40x (0.95NA) and a 20x (0.45NA) air objectives and a digital camera (Andor Zyla 4.2 sCMOS; Andor, Belfast, Ireland) (Bioimaging-DUTH facility). All system components were controlled by Andor IQ 3.6.5.

Quantification and determination of mean fluorescence intensity (MFI) from fluorescence microscopy images using confocal microscopy was performed with ImageJ software (ImageJ for Windows, Version 1.53k).

### Collagen measurement

The soluble collagen types (I–V) were determined using a Sircol Collagen Assay Kit. Briefly, PIFs were cultured in 6-well plates (Corning Inc.), treated with various agents and the culture supernatant was collected after 24 h, the time point at which the highest collagen production was observed. Cell debris was removed by centrifugation and the resulting supernatant was incubated overnight, at 4°C, with isolation and concentration reagent, provided with the kit. Τhe assay was therefore conducted according to the manufacturer’s protocol (CLRS1000; Biocolor), as previously described (13, 56).

### Multiplex cytokine measurement

The levels of multiple inflammatory cytokines were measured in serum and PIFs supernatants using the Human Inflammation Panel 1, LEGENDplex™ Multi-Analyte Flow Assay Kit (740809; Biolegend) in a CyFlow Cube 8 flow cytometer (Sysmex Partec, Germany), according to the manufacturer’s instructions as previously described (45).

### Immunohistochemistry (IHC-P), Masson’s trichrome and Immunofluorescence (IF) staining in tissue sections

To evaluate the presence of neutrophils in intestinal tissue biopsies, cross sections of 4 μm thickness, were stained using a mouse anti-human neutrophil elastase monoclonal antibody (NE; 1:50 dilution; clone NP57; DAKO). Serial cross sections, obtained from the same intestinal biopsies were further stained with Masson’s trichrome to identify the presence of intestinal fibrosis, following the manufacturer’s protocol (Masson-Goldner Trichrome kit, MGT-100T; Biognost). Samples were visualized under light microscopy (Nikon, model Eclipse E400) and images were captured using a Nikon Digital Camera (ACT-1 Nikon software).

Immunofluorescence staining was further performed in formalin-fixed paraffin-embedded biopsies, as previously described (57, 58). Briefly, nonspecific binding sites were blocked with 2% normal goat serum in 2% BSA-PBS. Paraffin-embedded tissue sections were deparaffinized and stained with either a rabbit anti-Smooth Muscle Actin monoclonal antibody (1:100 dilution; 701457; Invitrogen), or a rabbit anti-Vimentin monoclonal antibody (1:650; ab92547; Abcam), a mouse anti-KLF2 monoclonal antibody (6 μg/ml; MA5-24300; Thermo Fisher Scientific) or a mouse anti-IL-8 monoclonal antibody (15 μg/mL; MAB208-100; R&D systems. The next day and after extensive washes in PBS-1×, a Drop-n-Stain CF488A Goat anti-mouse IgG (H+L), highly cross-absorbed antibody (20956; Biotium) or a Drop-n-Stain CF594 Goat anti-rabbit IgG (H+L), highly cross-absorbed antibody (20955; Biotium) were utilized as the secondary antibodies. Tissues were mounted with DAPI (D9542, Sigma-Aldrich). Imaging was performed on a customized Andor Revolution Spinning Disc Confocal system with Yokogawa CSU-X1 (Yokogawa, Tokyo, Japan) confocal scan-head, build around an Olympus IX81 microscope (Olympus, Tokyo, Japan) mounted with a 40x (0.95NA) and a 20x (0.45NA) air objectives and a digital camera (Andor Zyla 4.2 sCMOS; Andor, Belfast, Ireland) (Bioimaging-DUTH facility). All system components were controlled by Andor IQ 3.6.5.

### *In-vitro* transwell migration assay (chemotaxis assay)

To evaluate neutrophil chemotaxis, low-serum complete DMEM medium suitable for the growth of intestinal fibroblasts (served as blank) and/or supernatant from HI, UC and CD PIFs were added to the lower chamber of a 24-well QCM^TM^ chemotaxis 3μm cell migration assay (ECM505; Merck Millipore), as described in the manufacturer’s instructions (59). In brief, freshly isolated human peripheral blood neutrophils (0,2-2 x 10^6^ cells/mL) were seeded in the upper well in low-serum complete DMEM. After 2.5 hours of incubation at 37°C and 5% CO_2_, migrating cells were quantified by measuring fluorescence using a Perkin Elmer fluorescence plate reader (Enspire, Waltham, MA, USA), set at excitation/emission wavelengths of 480/520 nm. Four independent experiments were performed, using neutrophils from a different healthy donor each time, in which mean fluorescence intensity was evaluated. As an inhibitor of neutrophil chemotaxis, a mouse anti-IL-8/CXCL8 monoclonal antibody with neutralizing effect (0,3 μg/mL; MAB208-100; R&D systems) was used, while an IgG rabbit polyclonal antibody (0,3 μg/mL; ab171870; Abcam) was used as a control in neutralization experiments.

## Supplemental Figures

**Figure S1.**
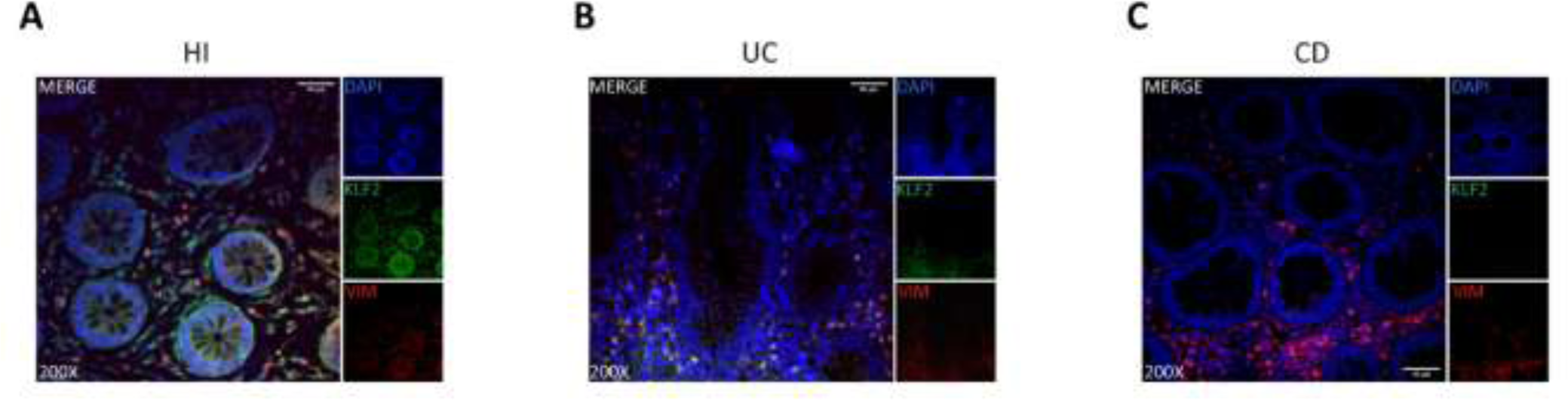
CD intestinal tissue fibroblasts are characterized by a KLF2 negative immunostaining. Immunofluorescence indicating KLF2 expression in **(A)** CD, **(B)** UC, **(C)** HI intestinal tissue fibroblasts (blue: DAPI, green: KLF2, red: Vimentin). One representative example out of four independent experiments is shown. Confocal microscopy. Magnification: 200x, Scale Bar: 40μm. CD, Crohn’s disease; HI, healthy individuals; KLF2, Kruppel-like Factor 2; UC, ulcerative colitis.

**Figure S2.**
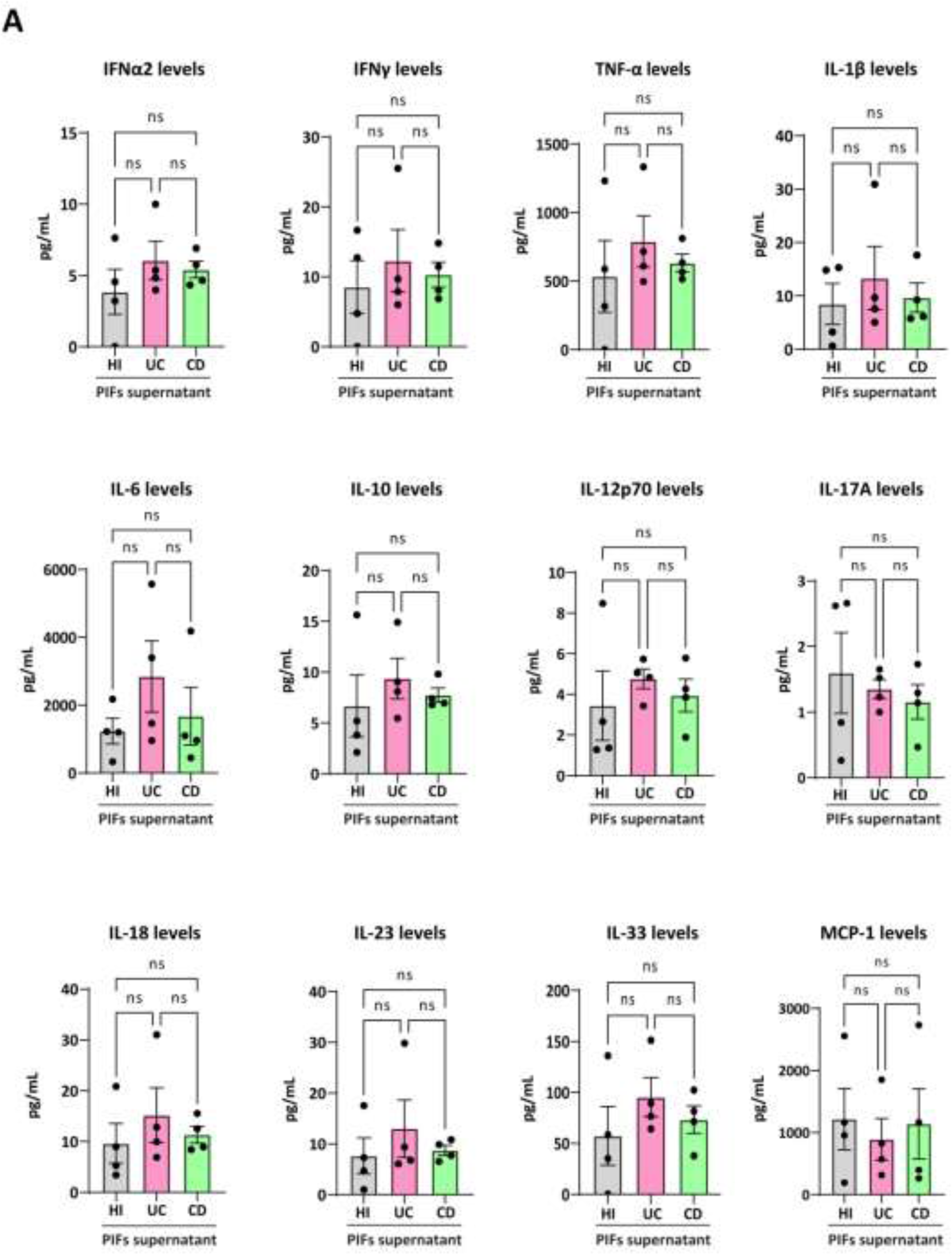
Inflammatory cytokines in supernatants of PIFs from IBD patients and HI. Levels of IL-1, IFNα2, IFNγ, MCP-1, TNF-α, IL-6, IL-10, IL-12p70, IL-17A, IL-18, IL-23 and IL-33 in the supernatants of ex-vivo isolated PIFs, as assessed by a bead-based multiplex flow cytometric assay. Nonparametric Kruskal-Wallis followed by Dunn’s multiple comparisons test was performed, *n=4*, n.s.: not significant. Data are expressed as mean ± SEM. CD, Crohn’s disease; HI, healthy individuals; MCP-1, Monocyte chemoattractant protein-1; PIFs, primary intestinal fibroblasts; TNF-α, tumor necrosis factor alpha; UC, ulcerative colitis.

**Figure S3.**
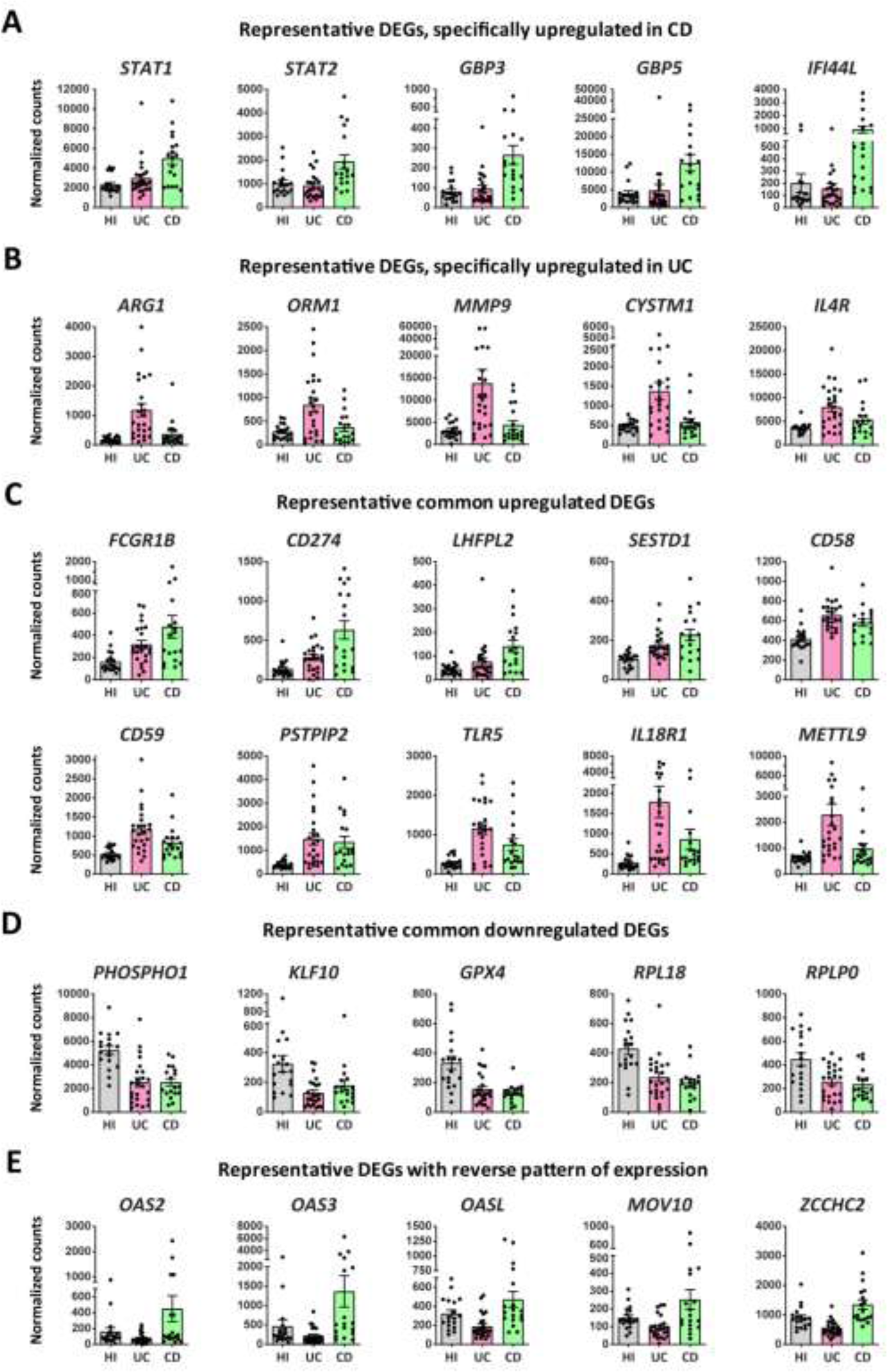
Representative differentially expressed genes in peripheral blood neutrophils isolated from patients with IBD and HI. Graphs of representative DEGs (baseMean >30 and FDR <0.05), specifically upregulated in CD **(A)** or UC **(B)**, commonly upregulated **(C)** or downregulated **(D)**, or showing a reverse expression pattern **(E)**, following RNA-Seq analysis of peripheral blood neutrophils from patients with CD (*n=18*) or UC (*n=24*). DEGs were identified following comparison with neutrophils isolated from healthy individuals (*n=18*). CD, Crohn’s disease; DEGs, differentially expressed genes; HI, healthy individuals; UC, ulcerative colitis.

**Figure S4.**
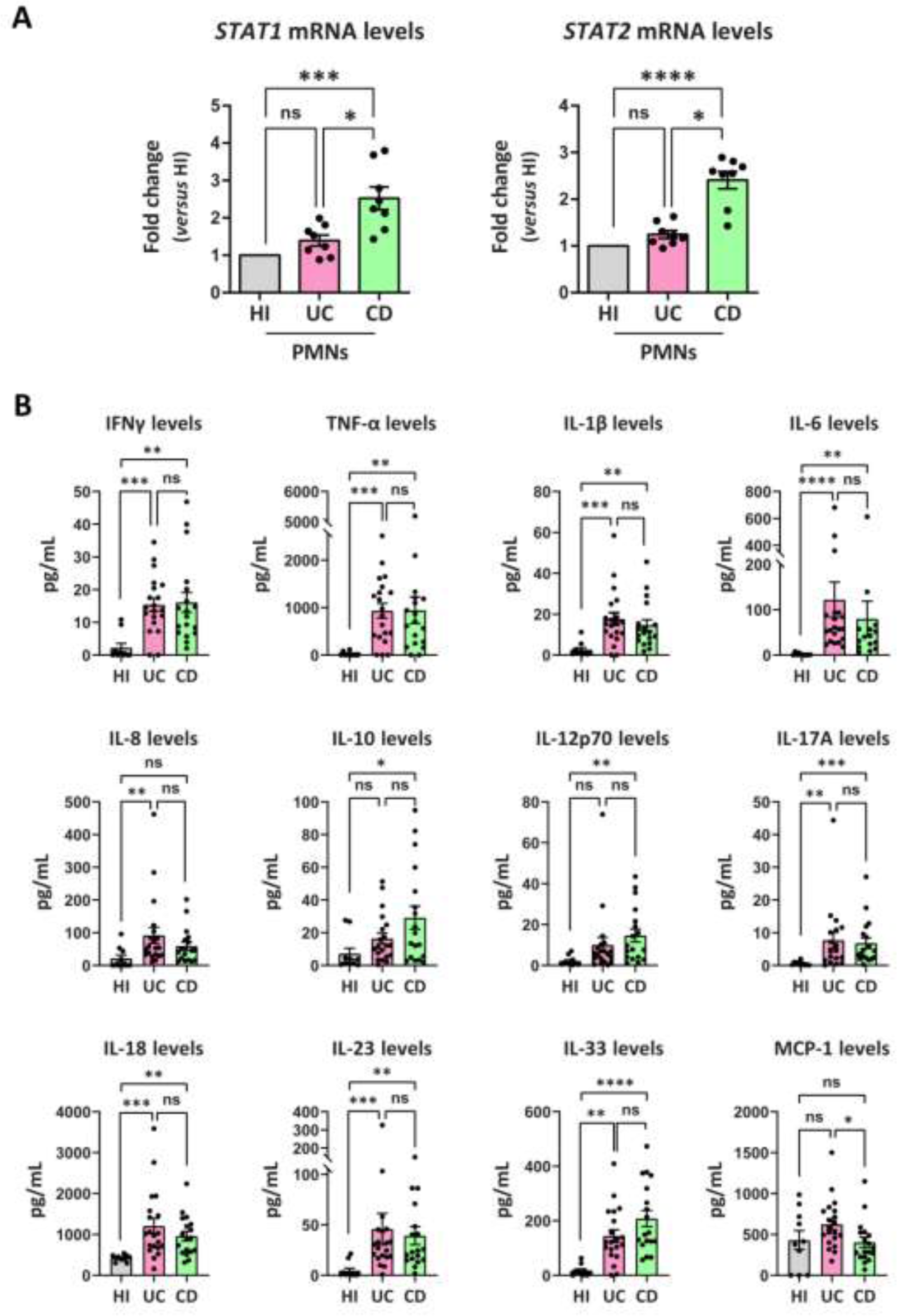
*STAT1* and *STAT2* mRNA levels in peripheral neutrophils and cytokine analysis in serum. **(A)** *STAT1* and *STAT2* mRNA levels of CD and UC patients, and HI, as assessed by qPCR (*n=8)*. **(B)** Serum levels of IL-1, IFNγ, MCP-1, TNF-α, IL-6, IL-8 IL-10, IL-12p70, IL-17A, IL-18, IL-23 and IL-33 as assessed by a bead-based multiplex flow cytometric assay in CD (*n=18*) and UC (*n=20*) patients, and HI (*n=10)*. **(A,B)** Nonparametric Kruskal-Wallis followed by Dunn’s multiple comparisons test was performed, *p<0,05, **p<0,01, ***p<0,001, ****p<0,0001, n.s.: not significant. Data are expressed as mean ± SEM. CD, Crohn’s disease; HI, healthy individuals; IFN, interferon; MCP-1, Monocyte chemoattractant protein-1; STAT, signal transducer and activator of transcription; TNF-α, tumor necrosis factor alpha; UC, Ulcerative colitis.

**Figure S5.**
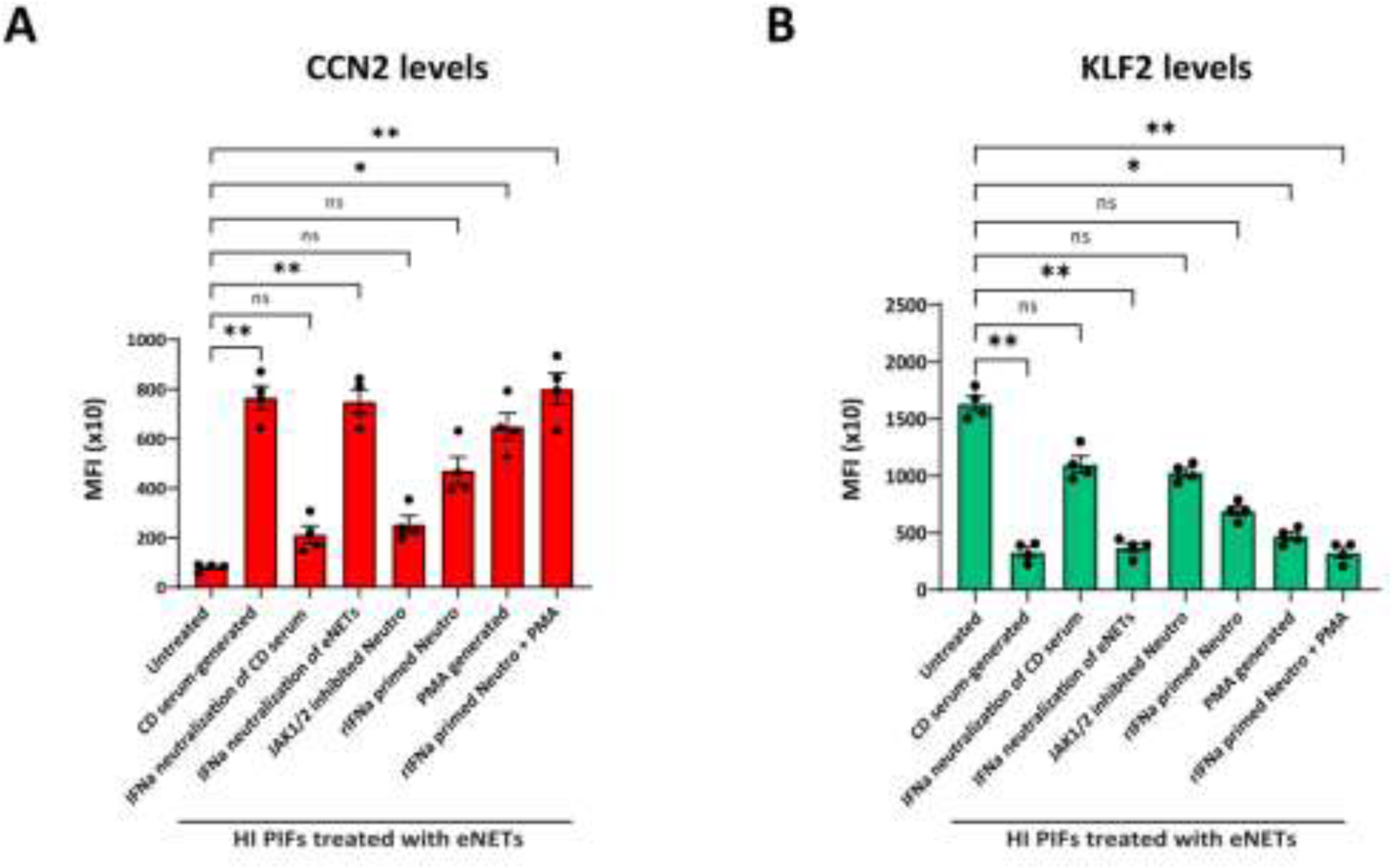
MFI quantification of HI PIFs treated with *in-vitro* generated eNETs. **(A)** Quantification of CCN2 and **(B)** KLF2 levels. **(A,B)** Nonparametric Kruskal-Wallis followed by Dunn’s multiple comparisons test, *n=6*, *p<0,05, **p<0,01, n.s.: not significant. Data are expressed as mean ± SEM. CCN2, cellular communication network factor 2; eNETs, enriched neutrophil extracellular traps; KLF2, Kruppel-like factor 2; MFI, mean fluorescence intensity; PMA, Phorbol-12-myristate-13-acetate.

**Figure S6.**
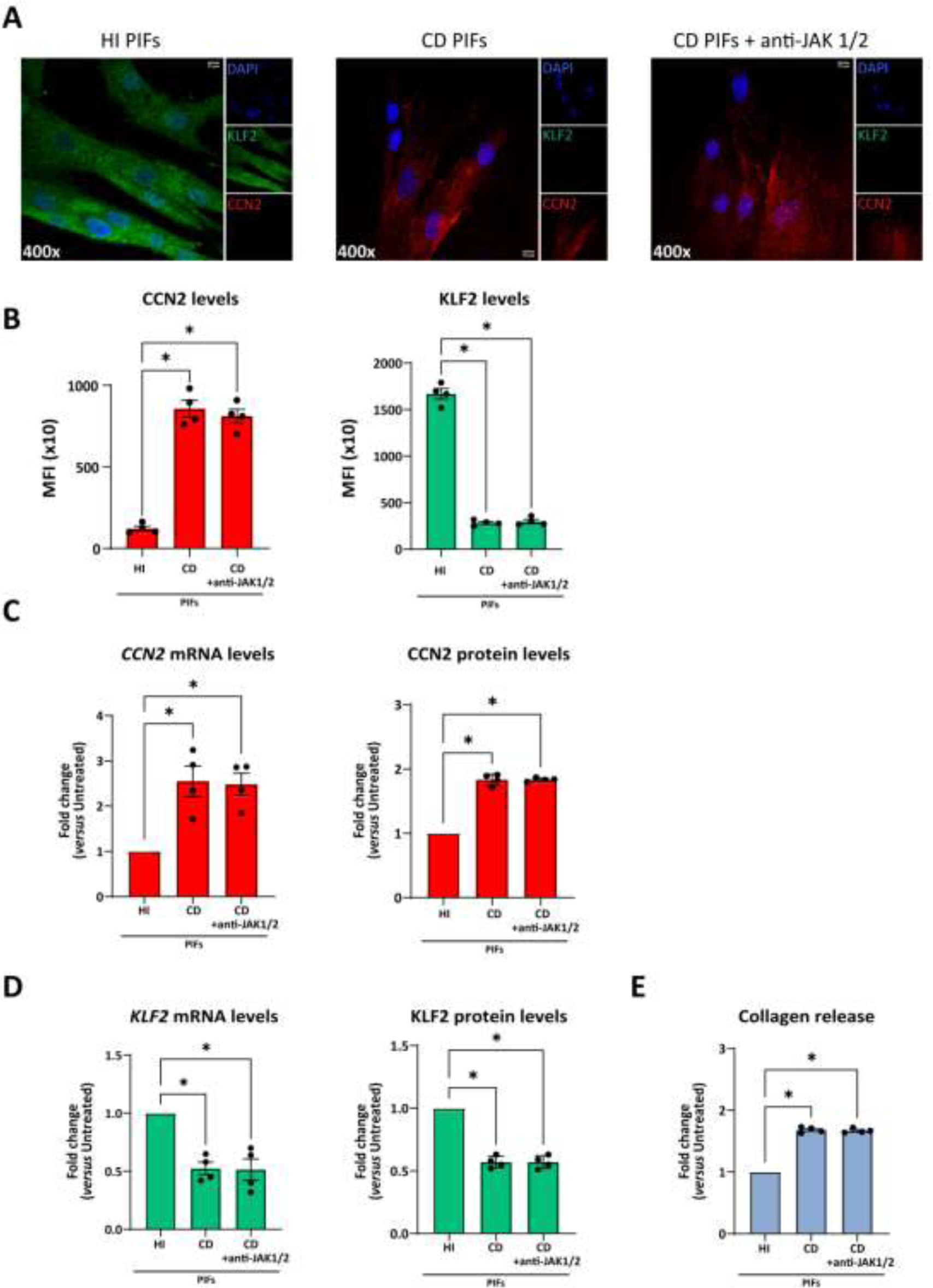
The fibrotic phenotype of CD PIFs is not reversed to normal by direct JAK-1/2 inhibition. JAK-1/2 inhibitor, baricitinib, was used to treat ex-vivo isolated CD PIFs. Untreated CD and HI PIFs were used as comparators. **(A)** KLF2 and CCN2 levels with immunofluorescence (blue: DAPI, green: KLF2, red: CCN2) and **(B)** corresponding MFI quantification. **(C)** CCN2 and **(D)** KLF2 mRNA and protein levels as assessed by qPCR and in-cell ELISA, respectively. **(E)** Collagen release assay in the above studies. **(A)** One representative example out of four independent experiments is shown. Confocal microscopy. Magnification: 400x, Scale Bar: 10μm **(B-E)** Nonparametric Kruskal-Wallis followed by Dunn’s multiple comparisons test was performed, *n=6*, *p<0,05, **p<0,01, ***p<0,001, ****p<0,0001, n.s.: not significant. Data are expressed as mean ± SEM. CCN2, cellular communication network factor 2; CD, Crohn’s disease; JAK, Janus Kinase; KLF2, Kruppel-like factor 2; MFI, mean fluorescence intensity; PIFs, primary intestinal fibroblasts.

**Figure S7.**
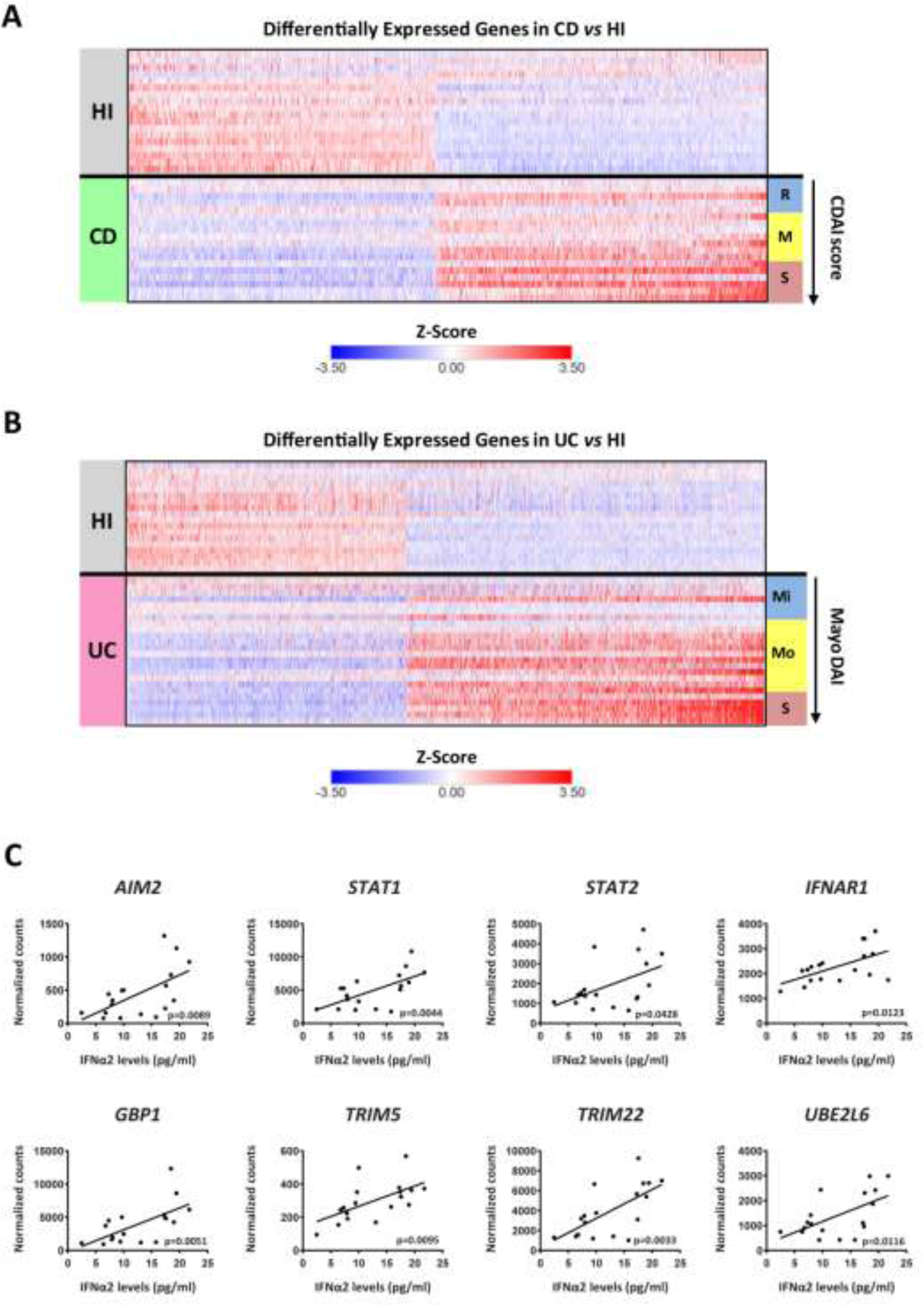
Transcriptomic alterations in peripheral blood neutrophils correlate with disease severity in IBD patients. **(A, B)** Heatmaps depicting the relative expression of all DEGs (baseMean > 30 and FDR < 0.05), as determined by RNA-Seq analysis of neutrophils isolated from healthy individuals (*n=18*) and CD (*n=18*), **A**) or UC patients (*n=24*), **B**). CDAI and Mayo DAI were used for assessing disease activity in CD and UC, respectively. **(C)** Correlation plots of the mRNA expression of key interferon signaling components, as determined by RNA-Seq analysis of CD neutrophils, *versus* serum IFNα2 levels. Simple linear regression was used to assess the relationship between normalized counts and IFNα2 levels. CD, Crohn’s disease; CDAI, Crohn’s disease activity index (R, remission, < 150; M, mild to moderate, 150-220; S, moderate to severe, > 220); FDR, false discovery rate; HI, healthy individuals; IBD, inflammatory bowel disease; Mayo DAI, Mayo score disease activity index (Mi, mild, 3-5; Mo, moderate, 6-10; S, severe, 11-12); UC, ulcerative colitis.

**Table S1.** Characteristics of healthy individuals and samples used (✓) in experiments.

| Sample | Sex/Age<br>(decade) | RNA-seq | Serum | PIFs | eNETs | Tissue | qPCR |
| --- | --- | --- | --- | --- | --- | --- | --- |
| HI1 | M/3rd | ✓ |  |  |  |  |  |
| HI2 | F/3rd | ✓ |  |  |  |  |  |
| HI3 | F/3rd | ✓ | ✓ |  | ✓ |  | ✓ |
| HI4 | M/7th | ✓ |  |  |  |  |  |
| HI5 | M/6th | ✓ |  |  |  |  |  |
| HI6 | M/5th | ✓ | ✓ |  |  |  | ✓ |
| HI7 | F/3rd | ✓ | ✓ |  |  |  |  |
| HI8 | M/3rd | ✓ | ✓ |  |  |  |  |
| HI9 | M/4th | ✓ | ✓ |  | ✓ |  | ✓ |
| HI10 | F/4th | ✓ |  |  |  |  |  |
| HI11 | F/6th | ✓ |  |  |  |  |  |
| HI12 | M/5th | ✓ | ✓ | ✓ | ✓ | ✓ | ✓ |
| HI13 | M/6th | ✓ | ✓ | ✓ | ✓ | ✓ | ✓ |
| HI14 | M/4th | ✓ | ✓ | ✓ | ✓ | ✓ | ✓ |
| HI15 | M/4th | ✓ |  |  |  |  |  |
| HI16 | F/4th | ✓ | ✓ | ✓ | ✓ | ✓ | ✓ |
| HI17 | M/4th | ✓ |  |  |  |  |  |
| HI18 | M/5th | ✓ | ✓ |  |  |  | ✓ |

**Table S2.** Characteristics of CD patients and samples used (✓) in experiments.

| Sample | Sex/<br>Age<br>(decade) | †Montreal<br>score | #CDAI | Treatment<br>at sampling | RNA-seq | Serum | PIFs | eNETs | Tissue | qPCR |
| --- | --- | --- | --- | --- | --- | --- | --- | --- | --- | --- |
| CD1 | M/3rd | A2,L3,B1 | 165 | Naïve | ✓ | ✓ |  |  |  |  |
| CD2 | M/3rd | A2,L1,B1 | 158 | Naïve | ✓ | ✓ |  |  |  |  |
| CD3 | M/2nd | A2,L1,B2 | 275 | Naïve | ✓ | ✓ | ✓ | ✓ | ✓ |  |
| CD4 | F/5th | A3,L1,B1 | 104 | Naïve | ✓ | ✓ |  |  |  |  |
| CD5 | F/5th | A3,L2,B1 | 89 | Naïve | ✓ | ✓ |  |  |  |  |
| CD6 | M/5th | A3,L3,B1 | 232 | Naïve | ✓ | ✓ |  | ✓ |  |  |
| CD7 | M/2nd | A2,L3,B3p | 295 | Naïve | ✓ | ✓ | ✓ | ✓ | ✓ |  |
| CD8 | F/5th | A3,L3,B1 | 176 | Naïve | ✓ | ✓ |  |  |  |  |
| CD9 | F/4th | A2,L1,B1 | 122 | Naïve | ✓ | ✓ |  |  |  |  |
| CD10 | F/4th | A2,L1,B1 | 232 | Naïve | ✓ | ✓ | ✓ | ✓ | ✓ |  |
| CD11 | F/3rd | A2,L1,B1 | 136 | Naïve | ✓ | ✓ |  |  |  |  |
| CD12 | M/3rd | A2,L3,B3 | 160 | Naïve | ✓ | ✓ |  |  |  |  |
| CD13 | M/5th | A2,L3,B3p | 161 | Naïve | ✓ | ✓ |  |  |  |  |
| CD14 | M/2nd | A2,L1,B2 | 182 | Naïve | ✓ | ✓ |  |  |  |  |
| CD15 | F/7th | A3,L1,B1 | 251 | Naïve | ✓ | ✓ |  | ✓ |  |  |
| CD16 | M/3rd | A2,L1,B2 | 274 | Naïve | ✓ | ✓ | ✓ | ✓ | ✓ |  |
| CD17 | M/4th | A2,L1,B2 | 190 | Naïve | ✓ | ✓ |  |  |  |  |
| CD18 | M/4th | A2,L2,B1 | 132 | Naïve | ✓ | ✓ |  |  |  |  |
| CD19 | M/4th | A2,L2,B3 | 288 | Naïve |  |  |  |  |  | ✓ |
| CD20 | M/2nd | A1,L1,B1 | 244 | Naïve |  |  |  |  |  | ✓ |
| CD21 | F/2nd | A2,L1,B2 | 210 | Naïve |  |  |  |  |  | ✓ |
| CD22 | M/5th | A3,L2,B1 | 207 | Naïve |  |  |  |  |  | ✓ |
| CD23 | F/4th | A2,L1,B1 | 190 | Naïve |  |  |  |  |  | ✓ |
| CD24 | M/8th | A3,L1,B1 | 210 | Naïve |  |  |  |  |  | ✓ |
| CD25 | M/5th | A3,L2,B3 | 272 | Naïve |  |  |  |  |  | ✓ |
| CD26 | F/2nd | A2,L2,B1 | 235 | Naïve |  |  |  |  |  | ✓ |

†Montreal score classifies Crohn's disease according to (A) age of disease onset, (L) location of disease and (B) disease behaviour. A1: Below 16 years; A2: between 17 and 40 years; A3: above 40 years. L1: terminal ileum; L2: colon; L3: ileocolonic. B1: non-stricturing, non-penetrating; B2: stricturing (inflammatory strictures); B3: penetrating; "p": perianal disease modifier.
#CDAI, Crohn's disease activity index. 0-150 points: Disease remission; 150-220: mildly to moderately active disease; Above 220: moderately to severely active disease.

**Table S3.** Characteristics of UC patients and samples used (✓) in experiments.

| Sample | Sex/Age (decade) | *Montreal Score | <sup>§</sup>Mayo-DAI | Treatment at sampling | RNA-seq | Serum | PIFs | eNETs | Tissue | qPCR |
| --- | --- | --- | --- | --- | --- | --- | --- | --- | --- | --- |
| UC1 | F/3rd | E3,S2 | 9 | Naïve | ✓ |  |  |  |  |  |
| UC2 | M/7th | E2,S2 | 4 | Naïve | ✓ | ✓ |  |  |  |  |
| UC3 | M/7th | E3,S3 | 11 | Naïve | ✓ | ✓ |  | ✓ |  |  |
| UC4 | M/3rd | E2,S2 | 6 | Naïve | ✓ |  |  |  |  |  |
| UC5 | M/6th | E2,S1 | 5 | Naïve | ✓ | ✓ |  |  |  |  |
| UC6 | M/5th | E3,S2 | 8 | Naïve | ✓ | ✓ |  |  |  |  |
| UC7 | M/6th | E2,S2 | 6 | Naïve | ✓ | ✓ |  |  |  |  |
| UC8 | M/2nd | E1,S1 | 3 | Naïve | ✓ | ✓ |  |  |  |  |
| UC9 | M/7th | E3,S1 | 5 | Naïve | ✓ | ✓ |  |  |  |  |
| UC10 | M/5th | E1,S2 | 7 | 5-ASA<sup>^</sup> | ✓ |  |  |  |  |  |
| UC11 | M/5th | E3,S1 | 6 | 5-ASA | ✓ | ✓ |  |  |  |  |
| UC12 | M/7th | E1,S1 | 4 | Naïve | ✓ | ✓ |  |  |  |  |
| UC13 | F/3rd | E2,S2 | 6 | Naïve | ✓ | ✓ |  |  |  |  |
| UC14 | M/8th | E3,S3 | 12 | Naïve | ✓ | ✓ | ✓ | ✓ | ✓ |  |
| UC15 | M/5th | E2,S3 | 11 | Naïve | ✓ | ✓ |  |  |  |  |
| UC16 | M/7th | E3,S2 | 9 | Naïve | ✓ | ✓ | ✓ | ✓ | ✓ |  |
| UC17 | M/7th | E3,S3 | 12 | Naïve | ✓ | ✓ | ✓ | ✓ | ✓ |  |
| UC18 | M/7th | E3,S1 | 4 | 5-ASA | ✓ | ✓ |  |  |  |  |
| UC19 | F/6th | E2,S1 | 5 | 5-ASA | ✓ |  |  |  |  |  |
| UC20 | M/7th | E3,S2 | 8 | Naïve | ✓ | ✓ |  | ✓ |  |  |
| UC21 | F/5th | E3,S3 | 11 | Naïve | ✓ | ✓ | ✓ | ✓ | ✓ |  |
| UC22 | M/6th | E3,S2 | 9 | 5-ASA | ✓ | ✓ |  |  |  |  |
| UC23 | F/9th | E3,S2 | 7 | 5-ASA | ✓ | ✓ |  |  |  |  |
| UC24 | M/4th | E2,S2 | 6 | 5-ASA | ✓ | ✓ |  |  |  |  |
| UC25 | F/5th | E3,S3 | 11 | Naïve |  |  |  |  |  | ✓ |
| UC26 | F/8th | E3,S3 | 11 | Naïve |  |  |  |  |  | ✓ |
| UC27 | M/4th | E3,S3 | 11 | Naïve |  |  |  |  |  | ✓ |
| UC28 | F/7th | E2,S2 | 9 | Naïve |  |  |  |  |  | ✓ |
| UC29 | M/2nd | E2,S2 | 7 | Naïve |  |  |  |  |  | ✓ |
| UC30 | M/4th | E2,S2 | 6 | Naïve |  |  |  |  |  | ✓ |
| UC31 | M/5th | E1,S2 | 6 | Naïve |  |  |  |  |  | ✓ |
| UC32 | M/9th | E3,S2 | 9 | Naïve |  |  |  |  |  | ✓ |

\*Montreal score classifies ulcerative colitis according to extend (E) and severity (S). E1: Ulcerative proctitis; E2: Left-sided colitis; E3: extensive ulcerative colitis (pancolitis). S0: remission, no symptoms of UC; S1: mild UC, $\leq 4$ bloody stools daily, lack of fever, pulse $< 90$ bpm, hemoglobin $> 105$ g/L, ESR $< 30$ mm/h; S2: moderate ulcerative colitis: $> 4$ -5 stools daily but with minimal signs of systemic toxicity; S3: severe ulcerative colitis: $\geq 6$ bloody stools daily, pulse $> 90$ bpm, temperatures $> 37.5^{\circ}\text{C}$ , hemoglobin $< 10.5$ g/dL, ESR $> 30$ mm/h
<sup>§</sup>Mayo-DAI, Mayo disease activity index. 0-2 points: Disease remission; 3-5 points: mildly active disease; 6-10 moderately active disease; 11-12 severely active disease
<sup>^</sup>5-ASA, 5-aminosalicylic acid; 5-ASA was discontinued at least 48 hours before sampling

**Table S4.**
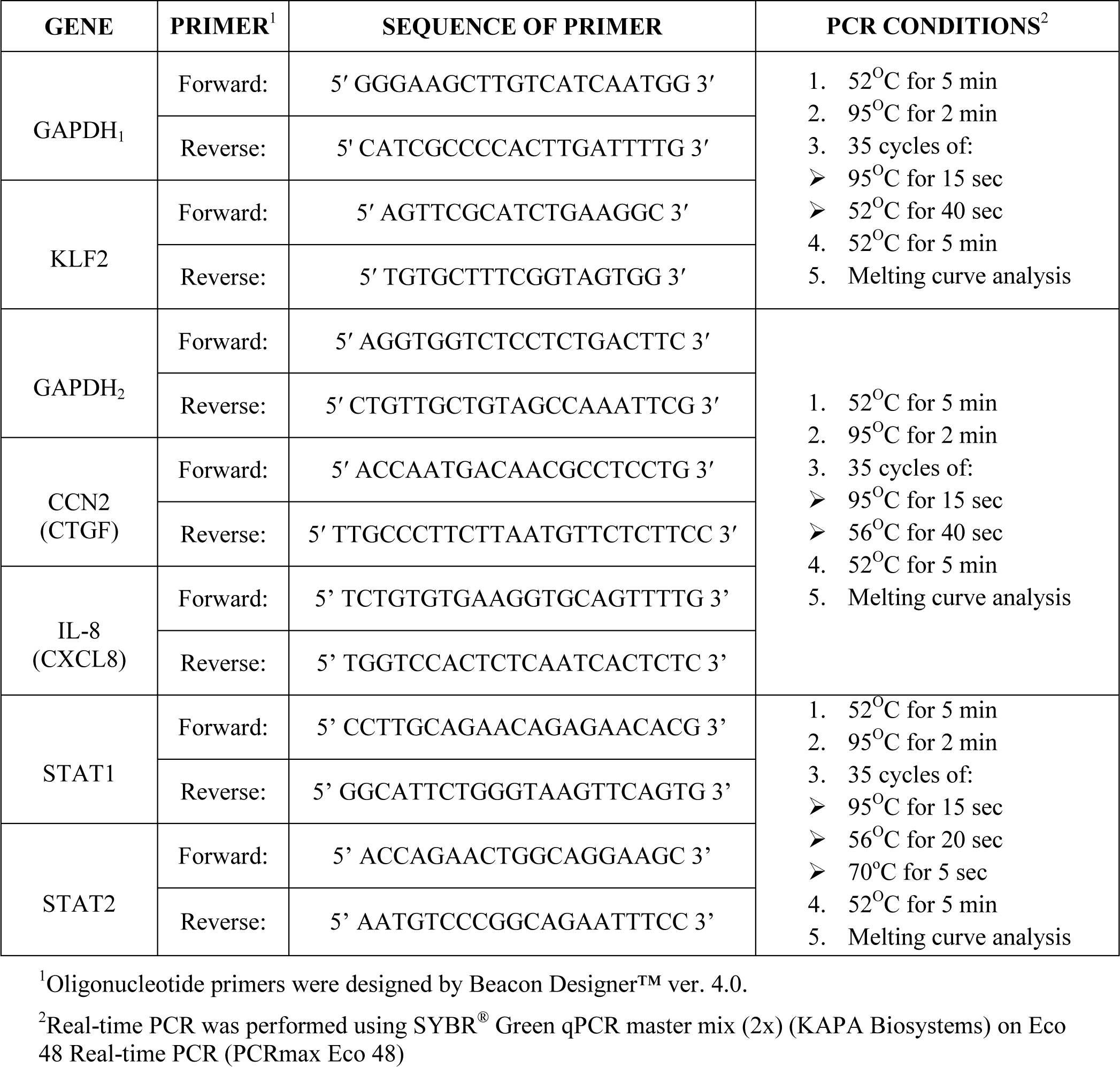
Sequence of Primers and real-time RT-PCR conditions.

